# Neuroinvasive Human Parvovirus 4 is associated with increased mortality in children: a multiyear retrospective observational study

**DOI:** 10.64898/2026.03.17.26348513

**Authors:** Deb Purna Keya, Apurba Rajib Malaker, Naito Kanon, Arif Mohammad Tanmoy, Saffat Reaz, Preonath Chondrow Dev, Hafizur Rahman, Lubana Tanvia, Anisur Rahman, Afroza Akter Tanni, Dipu Chandra Das, Anannya Barman Jui, Mirza Md. Ziaul Islam, Reaz Mobarak, Shamsun Nahar, Cristina Tato, ASM Nawshad Uddin Ahmed, Farhad Imam, Joseph L DeRisi, Samir K Saha, Yogesh Hooda, Senjuti Saha

## Abstract

**Background:** Infections of the central nervous system (CNS) in children remain a major cause of mortality and long-term disability globally, particularly in low- and middle-income countries (LMICs), where a high proportion of cases lack an identified pathogen. Sporadically, human parvovirus 4 (PARV4) has been detected in a small number of cerebrospinal fluid (CSF) from children with CNS infections, but its pathogenic role is unclear. We investigated the prevalence, clinical impact, and genomic characteristics of PARV4 in children with suspected meningitis.

**Methods:** We retrospectively analyzed CSF samples collected from children with WHO-defined suspected meningitis at the largest pediatric hospital in Bangladesh between 2015-2022. All samples underwent routine diagnostics, including bacterial culture and serological testing. Additional testing for PARV4 and parvovirus B19 was performed using qPCR of samples with >9 white blood cell (WBC)/µl followed by metagenomic sequencing of a subset. Clinical and laboratory data were extracted from patient records. Associations between PARV4 detection and mortality were assessed using logistic regression, adjusting for age, WBC count, and co-infections. Genomic and phylogenetic analyses were conducted on PARV4-positive samples.

**Findings:** Among 2,793 CSF samples with >9 WBC/µl, 526 (18·8%) were PARV4-positive. The median age of PARV4-positive cases was lower than that of PARV4-negative cases (4 vs 7 months, p<0·001). Co-infections were more common among PARV4-positive cases (49·6%) than PARV4-negative cases (16·4%). PARV4 positivity was independently associated with increased in-hospital mortality (adjusted odds ratio 2·09, 95%CI:1·46–2·96; p<0·001). Phylogenetic analysis indicated most strains belonged to genotype 2, with two sequences forming a distinct clade.

**Interpretation:** PARV4 is frequently detected in the CSF of children with suspected meningitis and is associated with increased in-hospital mortality. Its high prevalence, detection early in life, and frequent co-infection with other pathogens highlight the need to investigate PARV4 as an emerging CNS pathogen in LMICs.

## Introduction

Central nervous system (CNS) infections including meningitis remains associated with high morbidity and mortality worldwide.^1^ Globally, an estimated 10 million meningitis cases and 213,962 deaths occur each year, with 70% of these cases concentrated in low- and middle-income countries (LMICs).^2,3^ Survivors often experience long-term neurological sequelae, emphasizing the need for early diagnosis, pathogen-specific treatment, and prevention.^4^ Many pathogens including bacteria, viruses, fungi and parasites are known to cause CNS infections. In LMICs, where disease burden is the highest, pathogen detection rates remain particularly low: up to 80–85% of pediatric CNS infection cases may remain unexplained.^5,6^ This diagnostic gap undermines treatment, surveillance, and policy development for effective prevention strategies, including vaccine prioritization.

In recent years, the application of multiplex PCR panels and unbiased metagenomic sequencing has expanded our understanding of CNS infection etiology, leading to the detection of emerging or previously unrecognized viruses in cerebrospinal fluid (CSF), such as chikungunya virus, SARS-CoV-2, and parechoviruses.^7–9^ These discoveries suggest that additional understudied viruses may contribute to the burden of CNS infections in children, particularly in LMICs where surveillance is limited.

In recent years, human parvovirus 4 (PARV4 or tetraparvovirus) has surfaced as a virus of interest in CNS infections.^10,11^ Two independent studies from India reported PARV4 DNA in the CSF of children with suspected CNS infection (4 of 22 cases tested). A previous study from Bangladesh also detected PARV4 RNA in 3 of 25 CSF samples from suspected meningitis cases using unbiased RNA metagenomics.^9^ However, these findings, were limited by small sample sizes and lacked statistical association with clinical outcomes, leaving the clinical significance unresolved.

PARV4 is a small, non-enveloped, single-stranded DNA virus belonging to the *Parvoviridae* family. It was first identified in 2005 using sequence-independent PCR in an individual with febrile illness and hepatitis-like symptoms.^12^ It is one of only four parvoviruses known to infect humans, the others being parvovirus B19, human bocavirus, and adeno-associated viruses.^13^ The PARV4 genome is approximately 5·2 kb^14^, encoding two major open reading frames (ORFs).

While PARV4 is strongly associated with parenteral transmission, especially among people who inject drugs or those co-infected with HIV, hepatitis B virus (HBV), or hepatitis C virus (HCV), seroprevalence studies have also detected PARV4 IgG in individuals without these exposures, including healthy children, although the potential for serological cross reactivity to other viruses remains is unknown.^13–17^ Vertical transmission has also been documented in neonates born to IgM-positive mothers, with evidence of viremia at birth.^18^

Despite multiple reports of detection, the pathogenic role of PARV4 remains unclear. There is currently no *in vitro* culture system for PARV4, and whether it replicates autonomously is uncertain.^14^ Most infections appear to be asymptomatic, but PARV4 DNA has been detected coincident with respiratory, gastrointestinal, hepatic, and neurological symptoms.^11,12,19,20^ It has also been found in blood, bone marrow, lymphoid tissue, and liver.^13,21,22^

Overall, whether PARV4 contributes to disease, acts synergistically with other pathogens, or represents an innocuous passenger is not yet known. Its frequent co-detection with other viruses and association with young age raise questions about its tissue tropism, transmission dynamics, and relation to disease severity. In this study, we assess the prevalence, clinical associations, and genomic characteristics of PARV4 in a large collection of CSF samples from children with suspected meningitis, collected over eight years (2015-2022) at the largest pediatric hospital in Bangladesh. We evaluate associations with age, co-infections, and in-hospital mortality, and analyse genomic characteristics of circulating PARV4 strains.

## Methods

### Ethical considerations

The study protocol was approved by the Ethics Review Committee of the Bangladesh Institute of Child Health. Written informed consent was obtained from the parents or legal guardians of all participants prior to sample collection. All clinical data were de-identified prior to analysis.

### Study site and sample collection

Cerebrospinal fluid (CSF) samples were collected at Bangladesh Shishu Hospital and Institute (BSHI), formerly known as Dhaka Shishu Hospital, the largest tertiary pediatric hospital in the country, with 665 beds. BSHI and Child Health Research Foundation (CHRF) collaborate to conduct meningitis surveillance, which was part of the invasive bacterial vaccine-preventable disease (IB-VPD) surveillance.^9^

Lumbar puncture was performed as part of routine diagnostic evaluation for children presenting with suspected meningitis, at the discretion of the attending physician. Children were enrolled in the surveillance if their parent or legal guardian provided written informed consent, they met the WHO-defined case criteria for meningitis (Text S1; appendix p4) and had a CSF specimen collected as part of their clinical evaluation. Residual CSF specimens were stored at –80°C for long-term preservation and used for retrospective analyses in this study.

### Sample processing and screening

CSF specimens were cultured using standard microbiological methods and evaluated for meningitis causing pathogens (Text S2; appendix p4).

CSF samples with >9 WBC/µl, biobanked between 2015 and 2022, were screened for PARV4. We designed and optimized qPCR assays to detect PARV4 and *Erythroparvovirus* (B19) directly from whole CSF (Text S3; appendix p4). 16S Sanger sequencing was performed on culture-positive samples for which bacterial species could not be identified through standard microbiological methods (Text S4, appendix p5).

### Unbiased metagenomics and amplicon sequencing

Nucleic acid was extracted from 200 µl CSF samples using QIAamp DNA Mini kit (Qiagen, Germany) for DNA and Quick DNA/RNA Miniprep Plus kit (Zymo Research, USA) for RNA following the manufacturer’s instructions. Using previously optimized protocols, we used unbiased RNA and DNA metagenomics to detect PARV4 in the CSF samples and generate high coverage full genomes (Text S5; appendix p6).^9^

Near-complete genomes of PARV4 from unbiased metagenomics were used to establish a cost-effective amplicon sequencing protocol for PARV4 using PrimalScheme^23^ (Table S1; appendix p16, Figure S1; appendix p9) and following the protocol described in Text S6; appendix p6. One PARV4-confirmed CSF with a Ct (cycle threshold)-value <25 was randomly selected from each month between 2015-2022.

Consensus genomes of PARV4 from metagenomics and amplicon sequencing data were generated using CZ-ID viral consensus genome pipeline v3·5.0, an open source and cloud-based bioinformatics portal for analyzing NGS data (Text S6; appendix p6). We used 30 available sequences from Matthew *et al*^24^ to develop a genotyping scheme for PARV4 sequences using custom python scripts (Text S7; appendix p7).

### Genomic and phylogenetic analysis

To obtain multiple sequence alignment, all near complete genomes were aligned using MAFFT v7·490. Maximum likelihood tree (MLT) was constructed employing the generalized time-reversible model and a gamma distribution to model site-specific rate variation. 100 bootstrap replicates were used to assess MLT statistical support. We generated an outgroup rooted MLT (using MH215556·1 as the outgroup) with global sequences and visualized it.

We estimated genome-wide substitutions/year (Text S8; appendix p7, Table S5; appendix p26) and analyzed the mutation profile of PARV4 (Text S9, appendix p7).

### Clinical data collection and analysis

Demographic and clinical data were systematically collected from electronically stored health records for all cases. Descriptive analyses of the variables are presented as frequencies (percentages) or medians with interquartile ranges (IQR), as most variables exhibited non-normal distributions. Mann-Whitney U and Pearson chi-square tests were used to compare demographic and clinical characteristics between PARV4-positive and PARV4-negative groups. A two-proportion test was conducted to compare mortality rates between the two groups.

To further assess the association between PARV4 positivity and mortality, we performed both crude and adjusted logistic regression analyses. The adjusted model accounted for potential confounders, including age, WBC count, and co-infections, to provide a comprehensive understanding of the odds of mortality in PARV4-positive cases. All statistical tests were two-sided, with a significance level set at 5%.

## Results

### PARV4 prevalence, demographics, and clinical outcomes

Between 2015 and 2022, a total of 8,238 CSF samples were collected from pediatric patients presenting with suspected meningitis at BSHI (Figure 1). Among these, 3,235 samples had elevated WBC counts (>9 WBC/μl), of which 2,793 samples were preserved in the biobank. Samples were evenly distributed over the eight-year period, averaging 349 stored samples per year, except for 2020 when surveillance was interrupted due to COVID-19 lockdowns. All 2,793 stored CSF samples were tested for PARV4 using qPCR, with 526 (18·8%) testing positive for PARV4 DNA. 64·4% of the samples were collected within 72 hours of hospitalization (Table S2; appendix p19). Among PARV4-positive samples, Ct values were broadly distributed, with a median of 30·2 (IQR 24·4-33·9), with 12·9% of samples below Ct<20, and 30·0% of samples below Ct<25 (Figure S2; appendix p10).

**Figure 1.**
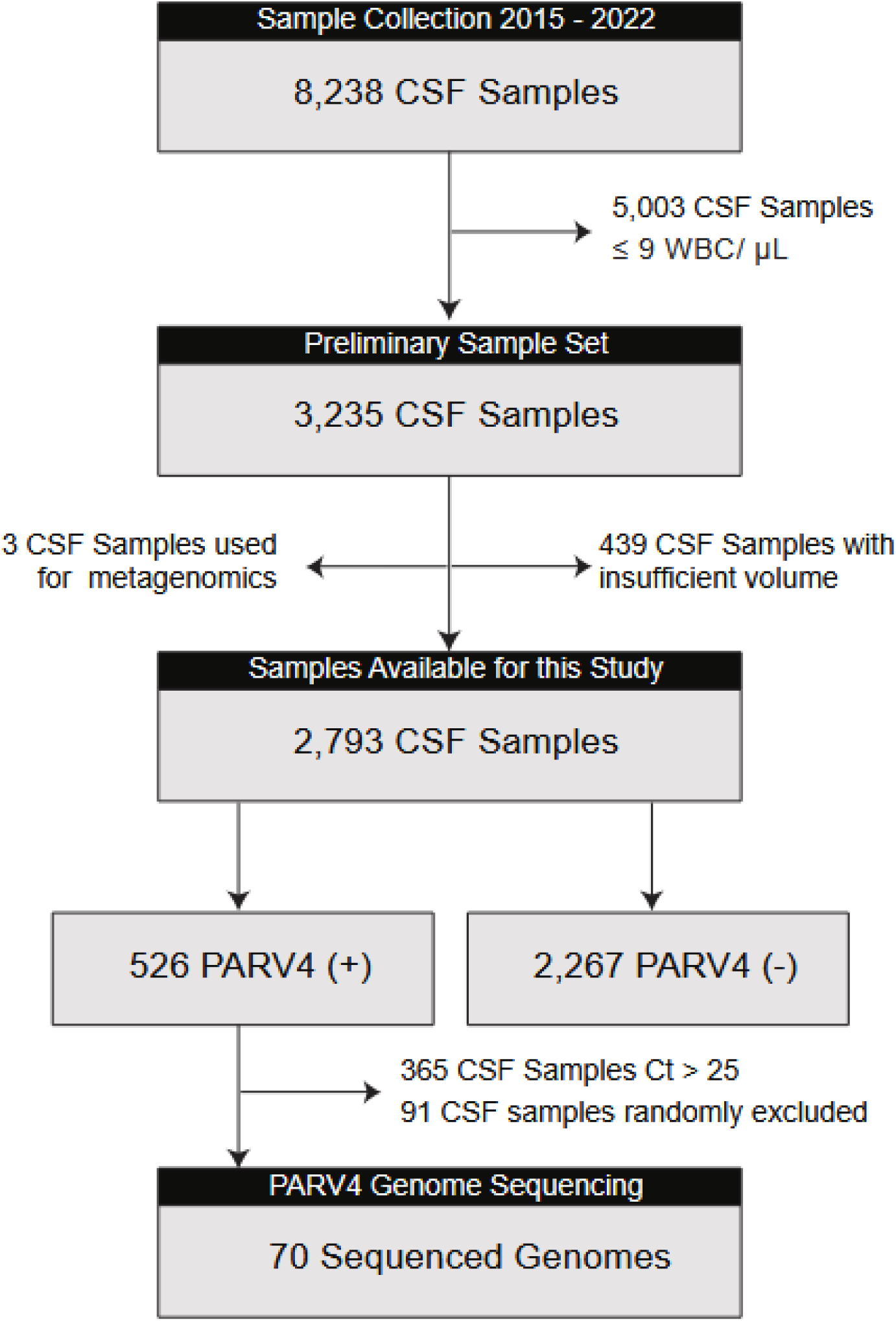
Flow diagram of CSF sample collection and PARV4 testing. This figure illustrates the process of sample collection from children under 5 years of age meeting the case definition of suspected meningitis at the Bangladesh Shishu Hospital and Institute. Samples were tested for PARV4, and eligible positive samples underwent genome sequencing.

The PARV4 positivity rate remained consistent over the study years, with the highest rate in 2021 at 31% and the lowest in 2016 at 11% (Figure S3A; appendix p11). Monthly prevalence analysis across the eight-year study period showed fluctuations in positivity rates but there appeared to be no consistent seasonal trend (Figure S3B; appendix p11). BSHI is the largest tertiary pediatric care hospital in Bangladesh, and cases included in this study came from different regions from across the country, with 24·7% of positive cases coming from the Dhaka district, where the hospital is located, and the remaining from other districts (Figure S4; appendix p12). Some areas had higher positivity rates than others, such as Barisal, Bhola, and Bogura, but this study was not designed or powered to assess statistical differences in spatial distribution.

The age distribution of PARV4-positive cases showed a peak in early infancy (Table 1, Figure 2A). The median age of PARV4-positive cases was 4 months (IQR 1-17 months), significantly younger than PARV4-negative cases whose median age was 7 months (IQR 2-30, p<0·001). Among the positive cases, 12·9% of the cases were in their first 2 weeks of life, 55·3% were <6 months, and 79·6% were <2 years of age. Sex distribution was similar in all groups, with approximate 60% of cases being male.

**Figure 2.**
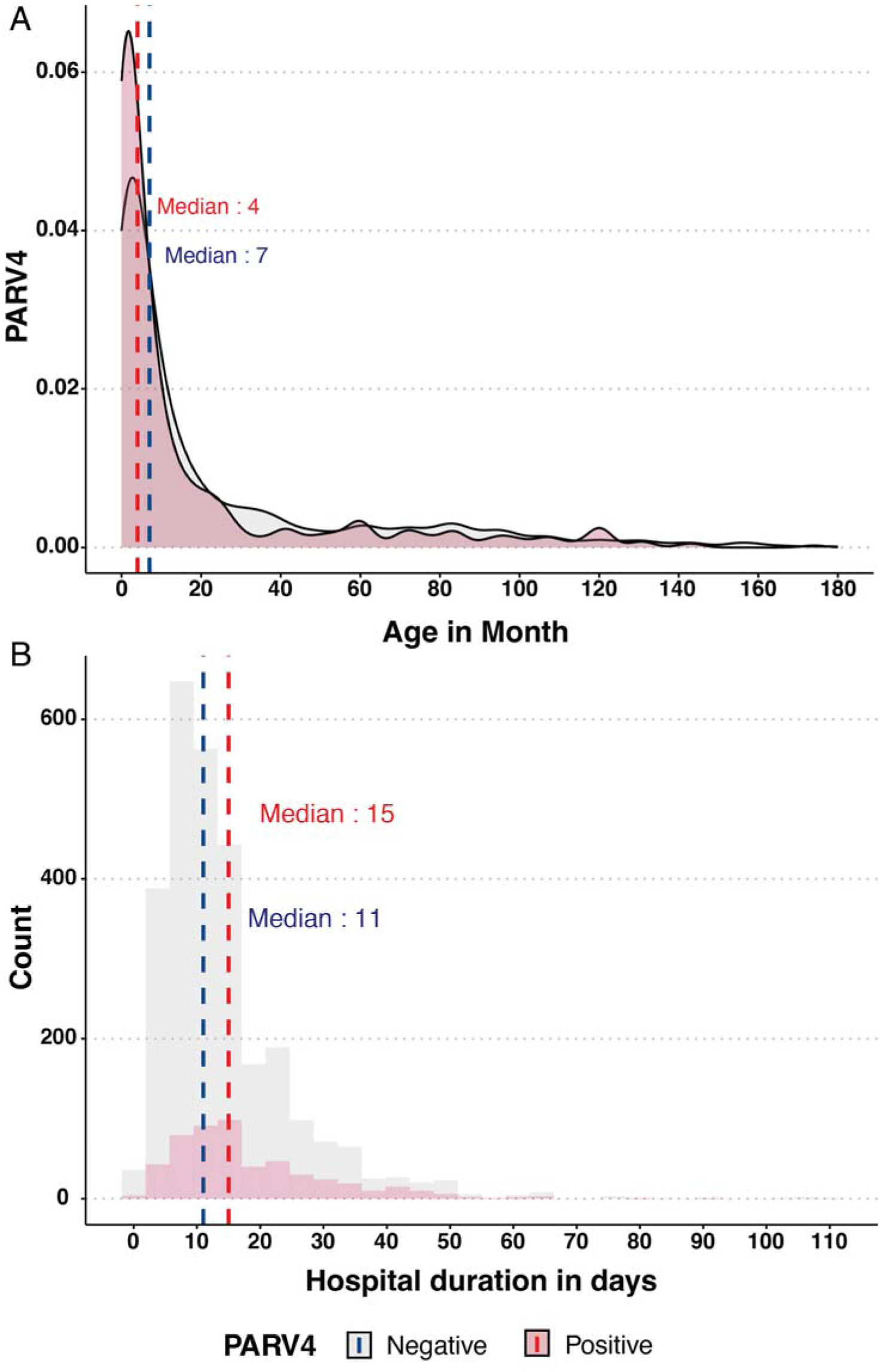
Age distribution and hospital duration of PARV4-positive and PARV4-negative cases. **(A)** Age distribution (months) of children with suspected meningitis according to PARV4 status; dashed lines indicate median ages. **(B)** Duration of hospital stay (days) by PARV4 status; dashed lines indicate median duration.

**Table 1.**
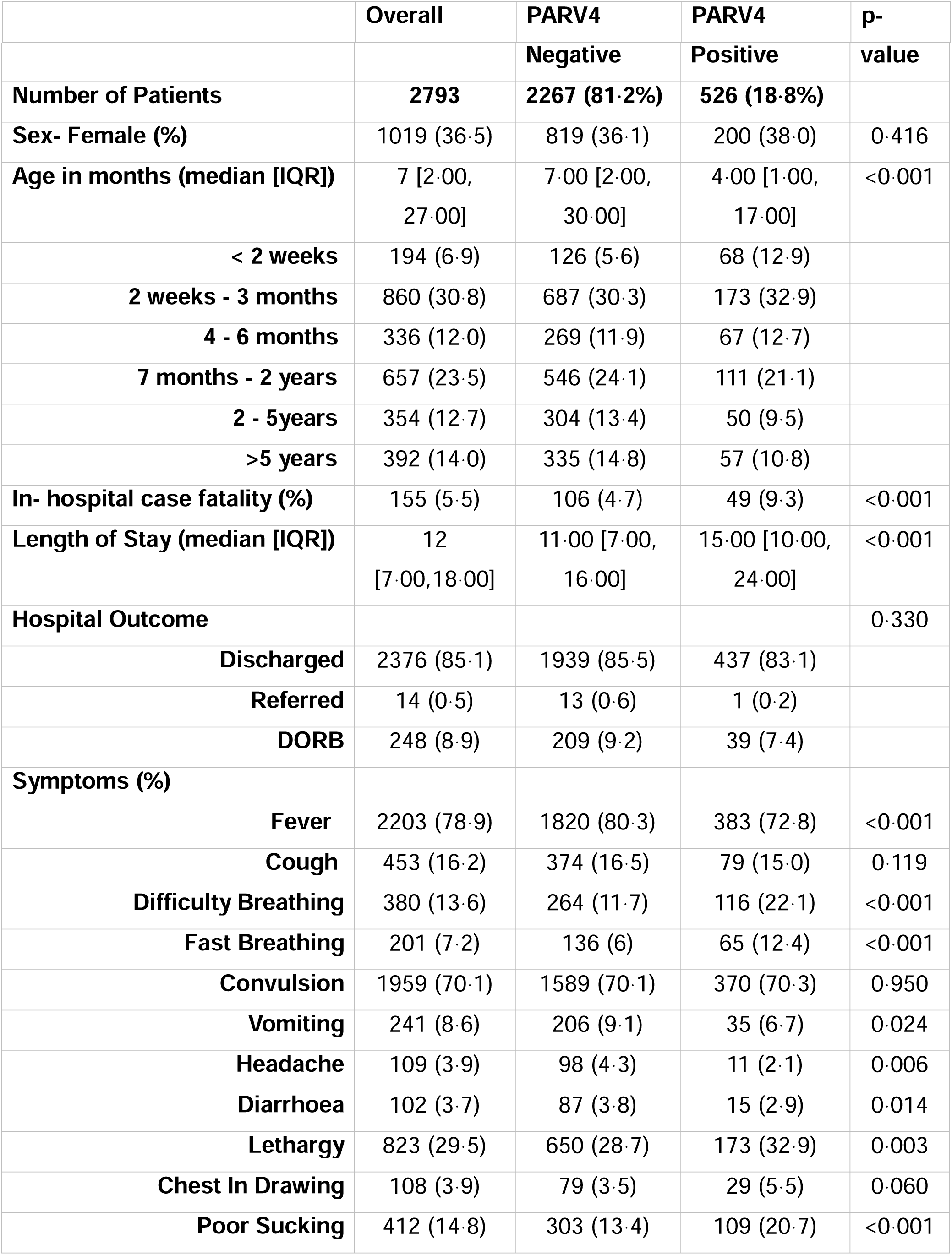
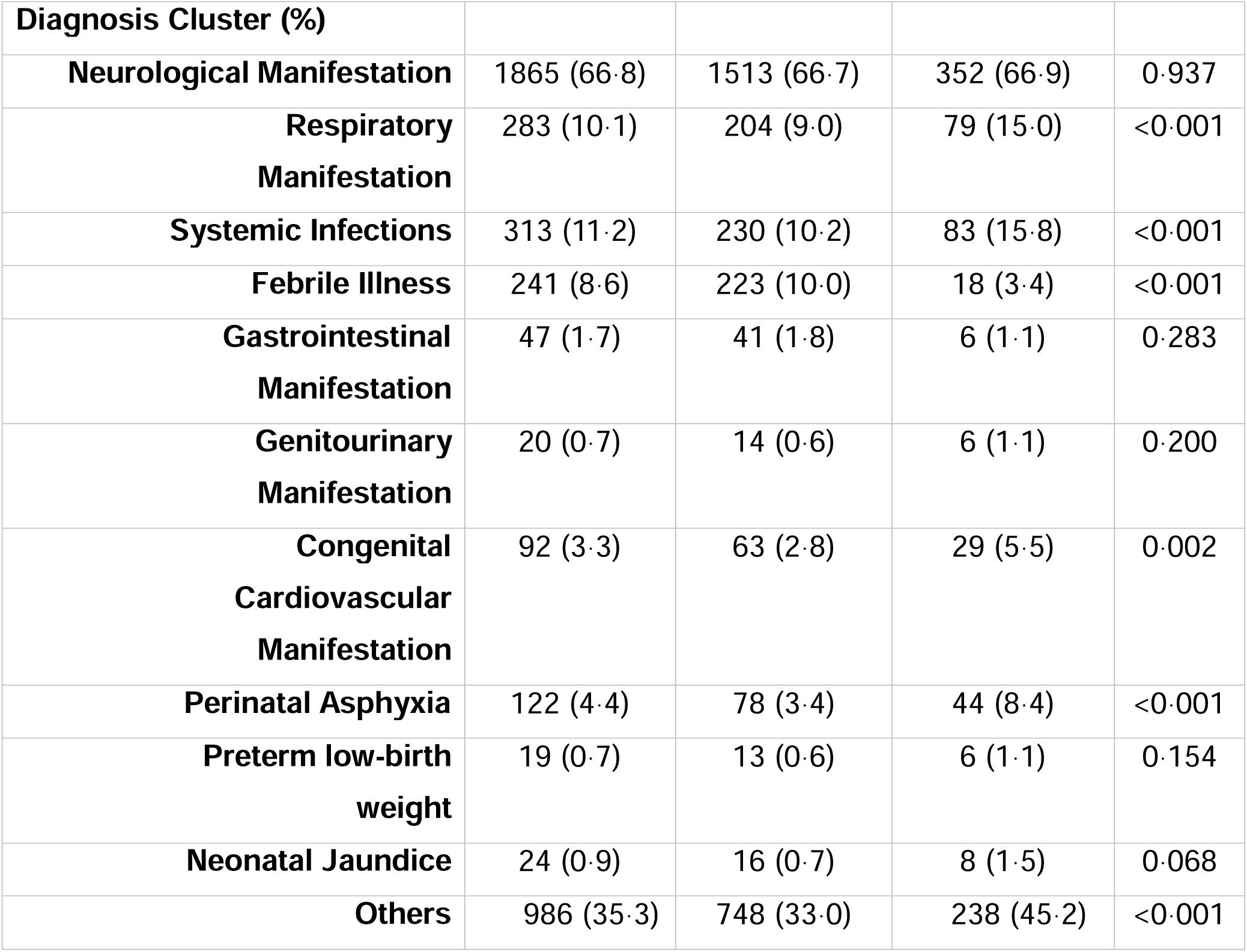
Clinical characteristics of cases (n = 2,793) admitted to study wards by PARV4 status. A list of diagnoses included in the “other” category is provided in Table S3; appendix p19). Data are n (%) or median [IQR] unless otherwise specified. p-values were calculated for positive vs. negative groups using the ranksum or chi-square test based on data type. DORB=discharge on risk bond.

|  | Overall | PARV4 | PARV4 | p- |
| --- | --- | --- | --- | --- |

|  |  | **Negative** | **Positive** | **value** |
| --- | --- | --- | --- | --- |
| **Number of Patients** | **2793** | **2267 (81.2%)** | **526 (18.8%)** |  |
| **Sex- Female (%)** | 1019 (36.5) | 819 (36.1) | 200 (38.0) | 0.416 |
| **Age in months (median [IQR])** | 7 [2.00, 27.00] | 7.00 [2.00, 30.00] | 4.00 [1.00, 17.00] | <0.001 |
| **< 2 weeks** | 194 (6.9) | 126 (5.6) | 68 (12.9) |  |
| **2 weeks - 3 months** | 860 (30.8) | 687 (30.3) | 173 (32.9) |  |
| **4 - 6 months** | 336 (12.0) | 269 (11.9) | 67 (12.7) |  |
| **7 months - 2 years** | 657 (23.5) | 546 (24.1) | 111 (21.1) |  |
| **2 - 5years** | 354 (12.7) | 304 (13.4) | 50 (9.5) |  |
| **>5 years** | 392 (14.0) | 335 (14.8) | 57 (10.8) |  |
| **In- hospital case fatality (%)** | 155 (5.5) | 106 (4.7) | 49 (9.3) | <0.001 |
| **Length of Stay (median [IQR])** | 12 [7.00,18.00] | 11.00 [7.00, 16.00] | 15.00 [10.00, 24.00] | <0.001 |
| **Hospital Outcome** |  |  |  | 0.330 |
| **Discharged** | 2376 (85.1) | 1939 (85.5) | 437 (83.1) |  |
| **Referred** | 14 (0.5) | 13 (0.6) | 1 (0.2) |  |
| **DORB** | 248 (8.9) | 209 (9.2) | 39 (7.4) |  |
| **Symptoms (%)** |  |  |  |  |
| **Fever** | 2203 (78.9) | 1820 (80.3) | 383 (72.8) | <0.001 |
| **Cough** | 453 (16.2) | 374 (16.5) | 79 (15.0) | 0.119 |
| **Difficulty Breathing** | 380 (13.6) | 264 (11.7) | 116 (22.1) | <0.001 |
| **Fast Breathing** | 201 (7.2) | 136 (6) | 65 (12.4) | <0.001 |
| **Convulsion** | 1959 (70.1) | 1589 (70.1) | 370 (70.3) | 0.950 |
| **Vomiting** | 241 (8.6) | 206 (9.1) | 35 (6.7) | 0.024 |
| **Headache** | 109 (3.9) | 98 (4.3) | 11 (2.1) | 0.006 |
| **Diarrhoea** | 102 (3.7) | 87 (3.8) | 15 (2.9) | 0.014 |
| **Lethargy** | 823 (29.5) | 650 (28.7) | 173 (32.9) | 0.003 |
| **Chest In Drawing** | 108 (3.9) | 79 (3.5) | 29 (5.5) | 0.060 |
| **Poor Sucking** | 412 (14.8) | 303 (13.4) | 109 (20.7) | <0.001 |

| **Diagnosis Cluster (%)** |  |  |  |  |
| --- | --- | --- | --- | --- |
| **Neurological Manifestation** | 1865 (66·8) | 1513 (66·7) | 352 (66·9) | 0·937 |
| **Respiratory Manifestation** | 283 (10·1) | 204 (9·0) | 79 (15·0) | <0·001 |
| **Systemic Infections** | 313 (11·2) | 230 (10·2) | 83 (15·8) | <0·001 |
| **Febrile Illness** | 241 (8·6) | 223 (10·0) | 18 (3·4) | <0·001 |
| **Gastrointestinal Manifestation** | 47 (1·7) | 41 (1·8) | 6 (1·1) | 0·283 |
| **Genitourinary Manifestation** | 20 (0·7) | 14 (0·6) | 6 (1·1) | 0·200 |
| **Congenital Cardiovascular Manifestation** | 92 (3·3) | 63 (2·8) | 29 (5·5) | 0·002 |
| **Perinatal Asphyxia** | 122 (4·4) | 78 (3·4) | 44 (8·4) | <0·001 |
| **Preterm low-birth weight** | 19 (0·7) | 13 (0·6) | 6 (1·1) | 0·154 |
| **Neonatal Jaundice** | 24 (0·9) | 16 (0·7) | 8 (1·5) | 0·068 |
| **Others** | 986 (35·3) | 748 (33·0) | 238 (45·2) | <0·001 |

Among the 2,793 patients evaluated, respiratory symptoms such as difficulty breathing (22·1% vs. 11·7%, p<0·001) and fast breathing (12·4% vs. 6·0%, p<0·001) were more frequent among PARV4-positive cases (Table 1). Poor sucking was also significantly higher in PARV4-positive cases (20·7% vs. 13·4%, p<0·001). Although neurological symptoms were common, with convulsions observed in 70·3% of PARV4-positive cases, this rate did not differ significantly in PARV4-negative cases (70·1%).

We also assessed differences in final diagnosis at death or end of hospitalization. Neurological manifestations were prevalent in both groups, affecting 66·9% of PARV4-positive cases, similar to 66·7% of PARV4-negative cases. However, respiratory manifestations and systemic infections were more common in PARV4-positive cases (15·0% and 15·8%, respectively) compared to PARV4-negative cases (9·0% and 10·2%, respectively; p < 0·001). The median WBC count for PARV4-positive cases was 50 cells/μl (IQR 20 - 210), which was the same (50 cells/μl) (IQR 20 - 170) for PARV4-negative cases (Figure S5; appendix p13).

The median length of hospital stay for PARV4-positive cases was significantly higher than for PARV4-negative cases (15 days vs 11 days, p<0·001; Figure 2B). There were no significant differences in the proportion of cases discharged, referred to other hospitals, or cases where patients left against medical advice (Table 1). However, the in-hospital case fatality rate among PARV4 positive cases was 9·3%, significantly higher than the 4·7% observed in PARV4-negative (p<0·001).

### Additional pathogens identified in PARV4-positive cases

All 2,793 CSF samples underwent bacteriological culture and immunochromatographic testing for *Streptococcus pneumoniae* (pneumococcus) antigen as part of routine diagnostics, and these results were available in the clinical records. In addition, in this study, we conduced qPCR for pneumococcus, *Neisseria meningitidis, Haemophilus influenzae*, and parvovirus B19 (B19). Among the 526 PARV4-positive cases, 262 (49·8%) were co-infected with at least one other pathogen, a rate significantly higher than that of the PARV4-negative cases, where in only 374 out of 2,267 (16·5%) cases another pathogen was detected (Table 2).

**Table 2.** Distribution of additional pathogens among PARV4-positive and PARV4-negative suspected meningitis cases. Organisms detected by culture, immunochromatographic test (ICT), or PCR in cerebrospinal fluid samples stratified by PARV4 status. Data are presented as number (%). B19 – erythroparvovirus B19, ICT – immunochromatographic test, PCR – polymerase chain reaction.

|  | PARV4 Positive (n=526) |  |  |  | PARV4 Negative (n=2267) |  |  |  |
| --- | --- | --- | --- | --- | --- | --- | --- | --- |
| Organism | Total | Culture | ICT | PCR | Total | Culture | ICT | PCR |
| B19 | 173<br>(32.9%) | 0 | 0 | 173 | 157<br>(6.9%) | 0 | 0 | 157 |
| *Streptococcus pneumoniae* | 62<br>(11.78%) | 2 | 59 | 1 | 136<br>(6.00%) | 5 | 130 | 1 |
| *Neisseria meningitidis* | 9 (1.71%) | 1 | 0 | 8 | 47<br>(2.07%) | 1 | 0 | 46 |
| *Haemophilus influenzae* | 5 (0.95%) | 0 | 0 | 5 | 11<br>(0.44%) | 0 | 0 | 11 |
| *Acinetobacter baumannii* | 3 (0.57%) | 3 | 0 | 0 | 3<br>(0.13%) | 3 | 0 | 0 |
| *Klebsiella pneumoniae* | 3 (0.57%) | 3 | 0 | 0 | 3<br>(0.13%) | 3 | 0 | 0 |
| *Escherichia coli* | 3 (0.57%) | 3 | 0 | 0 | 2<br>(0.09%) | 2 | 0 | 0 |
| *Elizabethkingia anopheles* | 2 (0.38%) | 2 | 0 | 0 | 1<br>(0.04%) | 1 | 0 | 0 |
| *Acinetobacter baumannii* | 1 (0.19%) | 1 | 0 | 0 | 3<br>(0.13%) | 3 | 0 | 0 |
| *Flavobacterium meningosepticum* | 1 (0.19%) | 1 | 0 | 0 | 2<br>(0.09%) | 2 | 0 | 0 |
| *Salmonella enterica*<br>serovar Typhimurium | 0 | 0 | 0 | 0 | 3<br>(0.13%) | 3 | 0 | 0 |
| *Desemzia incerta* | 0 | 0 | 0 | 0 | 1<br>(0.04%) | 1 | 0 | 0 |
| *Achromobacter xylosoxidans* | 0 | 0 | 0 | 0 | 2<br>(0.09%) | 2 | 0 | 0 |
| *Streptococcus agalactiae* | 0 | 0 | 0 | 0 | 1<br>(0.04%) | 1 | 0 | 0 |
| *Acinetobacter Iwoffii* | 0 | 0 | 0 | 0 | 1<br>(0.04%) | 1 | 0 | 0 |
| *Providencia alcalifaciens* | 0 | 0 | 0 | 0 | 1<br>(0.04%) | 1 | 0 | 0 |
| No organism detected | 264<br>(50.4%) | 0 | 0 | 0 | 1893<br>(83.6%) | 0 | 0 | 0 |

B19 was the only other virus tested for using PCR and was commonly detected in PARV4-positive cases (173 of 526 cases, 32·9%). In comparison, only 157 out of 2,267 PARV4-negative cases (6·9%) were B19-positive. Pneumococcus (the only bacteria whose presence is tested both through serology and culture) was the second most frequently detected organism in PARV4-positive cases, found in 62 of 526 cases (11·8%). This was almost double the detection rate in PARV4-negative cases, where 136 out of 2,267 cases (6·0%) tested positive for *Streptococcus pneumoniae*. Other bacterial pathogens like *Neisseria meningitidis* and *Haemophilus influenzae* were rarely detected in either PARV4 positive or negative cases.

### Association of PARV4 with in-hospital case fatality

We examined the odds of in-hospital mortality associated with PARV4 positivity while adjusting for demographic and clinical factors. Individuals testing positive for PARV4 had 2·09 times higher odds of mortality compared to PARV4-negative individuals (Figure 3). This association remained consistent when adjusted for age (aOR = 2·06) and for WBC count (aOR = 2·05) in separate models. After accounting for co-infection with only B19, the commonly detected co-pathogen, the odds of mortality in PARV4-positive cases remained 2·13 times higher than in the negative group, regardless of B19 co-infection status.

**Figure 3.**
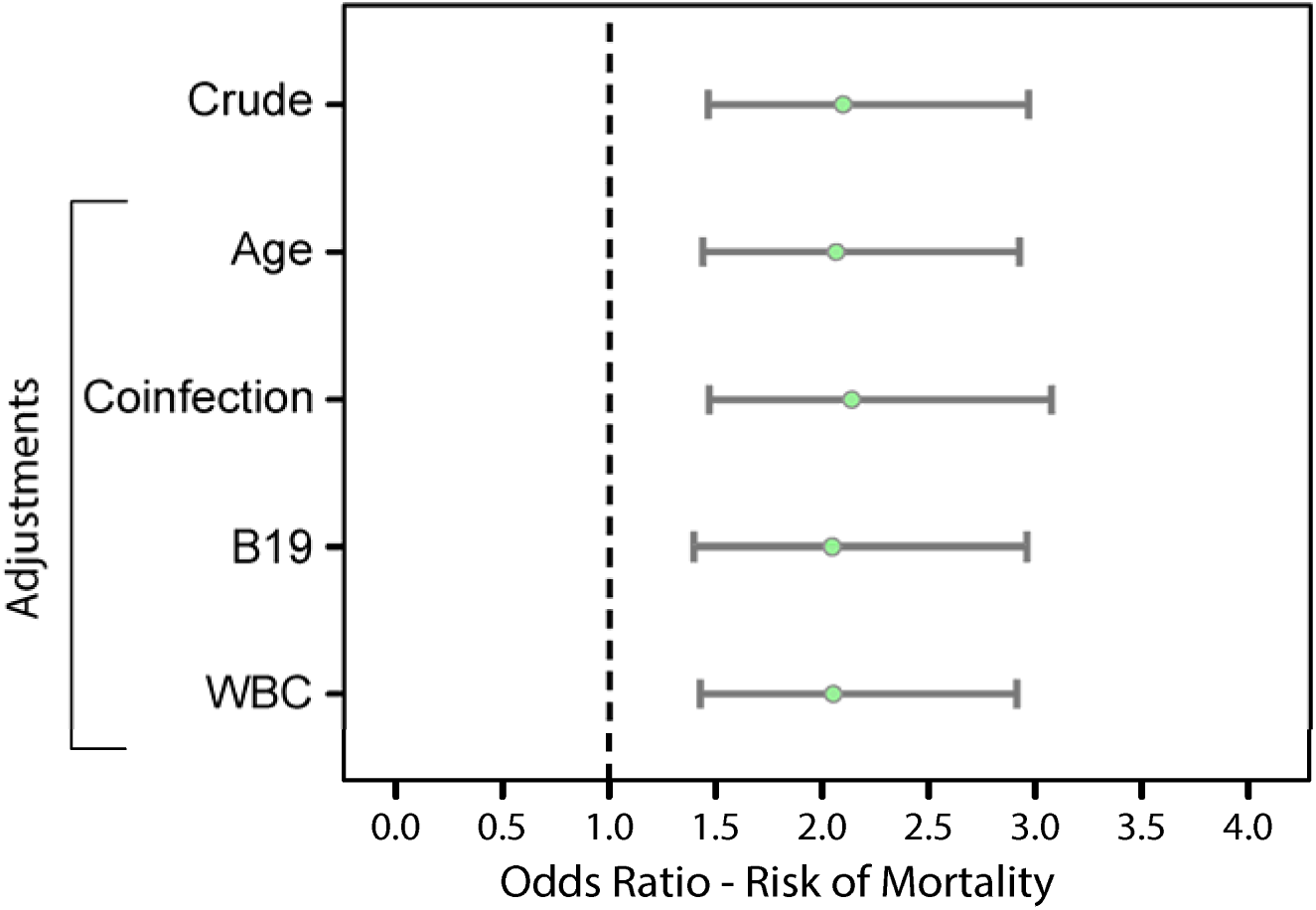
Association between PARV4 positivity and in-hospital case fatality. Odds ratios (ORs) for in-hospital case fatality associated with PARV4 positivity under crude and adjusted models. Models were adjusted for age, other pathogens detected, erythroparvovirus B19, and white blood cell (WBC) count. Points indicate ORs and horizontal lines show 95% CIs; the vertical line at OR=1 indicates no association.

### Genomic analysis of PARV4

A total of 73 samples underwent genome sequencing, including 20 sequenced using unbiased metagenomics and 53 using amplicon sequencing. Of these, 70 samples yielded sufficient data for whole genome assembly and further analysis. The coverage depth across amplicon-based sequences ranged from 1694x-31248·9x with 98% samples having coverage depth more than 1694x. In these samples, over 97% of the genome met thresholds for calling consensus bases. An overview of the consensus genomes with coverage depth is given in Table S4; appendix p24.

Bayesian analysis of Bangladeshi PARV4 population (n=70) estimated a genome-wide substitution rate of 0·5911475 substitutions per year (Table S5; appendix p26). This rate represents the average number of nucleotide changes accumulating in the PARV4 genome annually in Bangladesh. We then analysed the mutation profile of 66 PARV4 genomes which had complete ORF1 and 2 and used a local genome (CHRF_CSF_0090, accession number: ERS29405649) from 2015 as a reference. Non-synonymous (NS) mutation profiling revealed region-specific variability in the frequency of mutations (Figure 4A, Figure S6; appendix p14). ORF1 coding for NS1 exhibited sparse distribution of NS substitutions. In ORF2, coding for VP1 and VP2 structural proteins, NS substitutions were concentrated towards the unstructured VP1 region (non-VP2 region of VP1) with over 70% samples having amino acid substitutions at various regions between aa 913 and aa 1125. PARV4 showed limited variation along the capsid forming VP2 with 77·6% of samples showing substitutions in variable regions-II (VR-II) and 37·3% in VR-VIII.

**Figure 4.**
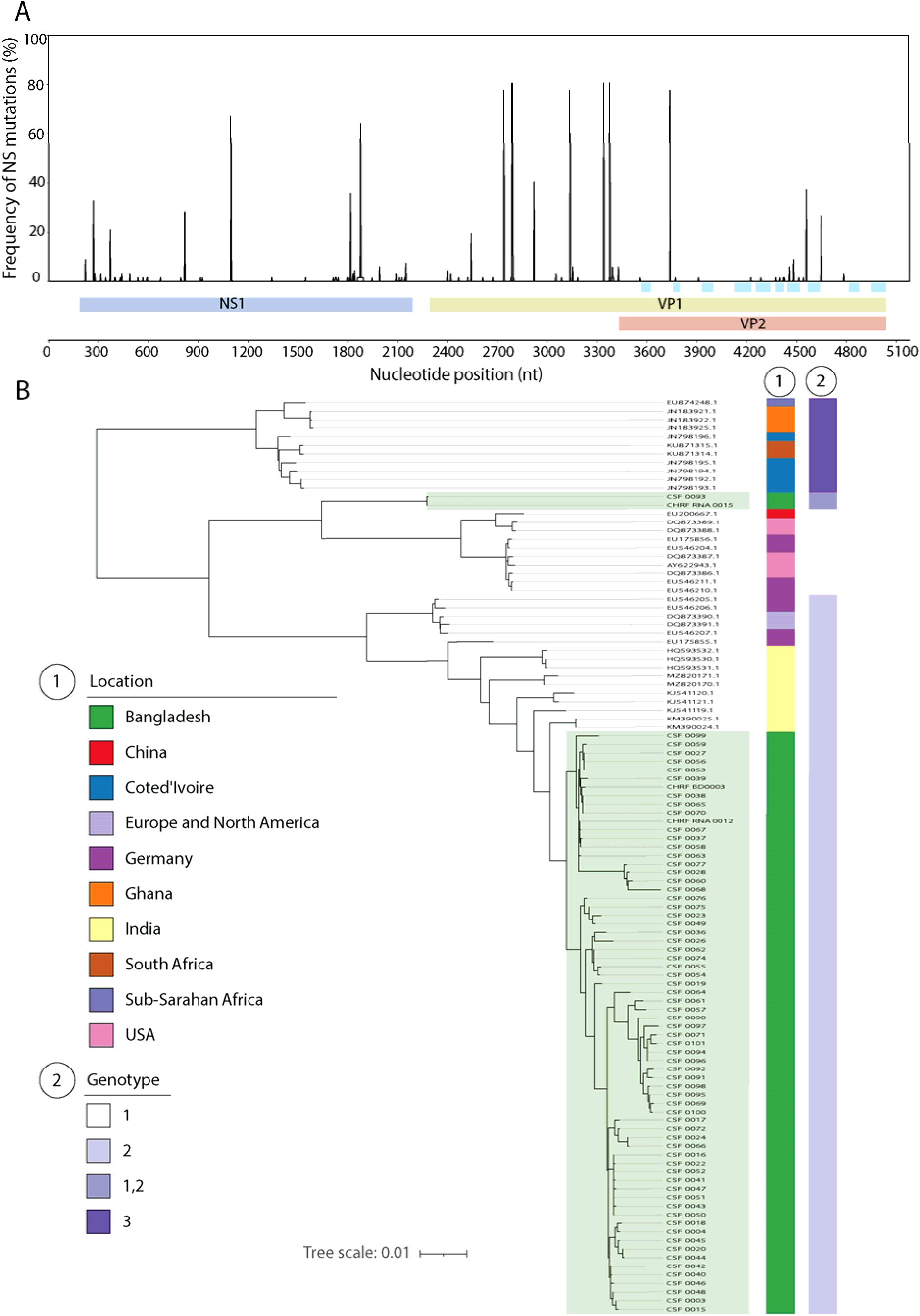
Genomic variation and phylogenetic analysis of PARV4. **(A)** Frequency of non-synonymous mutations across the PARV4 genome. The x-axis shows nucleotide position with annotated coding regions (NS1, VP1 and VP2); y-axis indicates the proportion of isolates with NS mutations at each position. Variable regions (VR-I to VR-IX and the HI loop) within VP2 region are highlighted in blue. **(B)** Maximum-likelihood phylogenetic tree of PARV4 genomes generated in this study together with publicly available global sequences. Outer annotations indicate geographic origin and genotype. The scale bar represents substitutions per site.

Phylogenetic analysis of the 70 assembled genomes and 37 publicly available PARV4 sequences showed that most sequences from Bangladesh clustered within genotype 2, aligning closely with strains previously identified in India (Figure 4B). Two sequences formed a distinct clade between genotypes 1 and 2, suggesting a potential novel genotype within Bangladesh (we term genotype 1,2).

## Discussion

With significant positive associations between PARV4 positivity, co-infections, and in-hospital case fatality, this study generates evidence that PARV4 is neuroinvasive and could be significant contributor to CNS infections particularly in settings where rates of idiopathic meningitis continue to be high.

We detected PARV4 in 18·8% of CSF samples from suspected meningitis cases. This rate aligns with the two reports from India - one detected PARV4 DNA in 2 of 10 CSF samples from children with acute CNS infection and another detected in 2 of 12.^10,11^ Our data also revealed a higher prevalence of PARV4 among children under 1 year of age, with a significantly lower median age of 4 months in PARV4-positive cases compared to PARV4-negative cases (7 months). 13% of the positive cases were in babies younger than 2 weeks. This trend may indicate increased susceptibility to PARV4 infection in early life with potential vertical transmission. In a study from Taiwan, placental transmission of PARV4 was reported in neonates who presented with viremia and were born to IgM-positive mothers, suggesting the virus may be transmitted vertically, in line with our findings.^18^

Almost half of the PARV4-positive cases had at least one additional pathogen detected in the CSF. The most common co-infection was with parvovirus B19 (only virus tested for), a related but distinct member of the same viral family. Parvovirus B19 was detected in a third of PARV4-positive cases compared to only 6·97% in PARV4-negative cases. Several previous studies have also reported strong association of PARV4 with other viruses like HBV, HCV and HIV.^13,16,17^ In addition to B19, *Streptococcus pneumoniae* (SPN) was the second most frequently detected co-infecting pathogen in PARV4-positive cases. While the immunochromatographic test used for detection of pneumococcus in the CSF is highly sensitive compared to only culture used for other bacteria,^25^ pneumococcal antigen was present in nearly double the rate in PARV4-positive cases compared to negative cases. Given that pneumococcus is a leading cause of bacterial meningitis, co-infection with PARV4 could complicate clinical management, particularly if PARV4 contributes to atypical presentations or immune modulation in infected individuals. Although co-infections with *Neisseria meningitidis* and *Haemophilus influenzae* were less common, overall higher rates of co-infections may suggest that PARV4 increases susceptibility to other CNS pathogens or causes secondary infections upon infection with other pathogens that impact clinical outcomes.

A key finding of this study is the association between PARV4 positivity and increased in-hospital mortality. Nearly 10% of PARV4-positive cases died during admission, and compared with PARV4-negative cases, PARV4-positive children had approximately twice the odds of death. This association remained after adjustment for age, white blood cell count, and detection of other pathogens. The relatively low cycle-threshold values observed, together with prior detection of PARV4 RNA in unbiased metagenomic studies^9^, suggest the likely possibility of active viral replication within the CNS, and potential neurotropism. Given that infants comprised of the majority of PARV4-positive cases, the observed mortality underscores the potential clinical relevance of PARV4 detection in this vulnerable population.

Beyond the differences in mortality, respiratory manifestations and systemic infections were more frequently observed among PARV4-positive cases than PARV4-negative cases. These findings suggest that PARV4-associated illness may have distinct clinical features compared to other pediatric CNS infections. Detection of PARV4 in respiratory samples from children with respiratory illness in Ghana and India has also been reported^19,26^, raising the possibility of respiratory involvement and potential respiratory transmission, similar to B19. However, further studies are needed to clarify transmission dynamics and clinical pathogenesis.

Bayesian phylogenetic analysis of the Bangladeshi PARV4 genomes revealed a genome-wide substitution rate of 0·5911475 substitutions per year, consistent with those observed in related human parvoviruses, which generally exhibit substitution rates between ∼1x10^-5^ and 1x10^-4^ substitutions per site per year.^27^ The unique N-terminal unstructured region of VP1 in PARV4 exhibited the presence of several NS mutations, suggesting a potential site for positive selection. In B19, this region of VP1 has been demonstrated to be important during the internalization of the virus in the host cell and was found to be present on the exterior side of capsid which could potentially enable interaction with their host for infection and to elicit immune response.^28,29^ The role of this region of VP1 in PARV4 pathogenicity remains unknown.

Phylogenetic analysis of 70 PARV4 genomes showed that most isolates clustered within genotype 2. This aligns with previous reports showing that the three PARV4 genotypes classified to date have varying geographic distributions: genotypes 1 and 2 are prevalent in Europe, North America, and Asia, while genotype 3 is common in sub-Saharan Africa.^13,17,30^ The PARV4 genomes sequenced from CSF samples in India also belong to genotype 2.^10^ Notably, two sequences formed a distinct clade positioned between genotypes 1 and 2, indicating the circulation of a distinct genotype within Bangladesh, which we term 1,2.

The findings of this study must be interpreted within the context of several limitations. First, our study was conducted at a single tertiary care hospital, potentially limiting the generalizability of the results to other settings in Bangladesh or other LMICs. However, the large dataset of over 2,000 samples collected over eight years from patients across the country suggests that our findings are likely to reflect broader trends and are not isolated from a single institution. Second, the cross-sectional design of the study restricts our ability to establish causality between PARV4 infection and clinical outcomes, including mortality. Although logistic regression analysis suggests a strong association, prospective and longitudinal studies, paired with laboratory studies with ex-vivo models are needed to confirm the role of PARV4 in disease causality, progression and outcomes. Finally, our detection methods relied on PCR and metagenomic sequencing, which, while sensitive, may not fully distinguish between active infection and chronic viremia. Future studies incorporating viral load quantification or markers of active infection could help determine whether PARV4 presence directly contributes to CNS pathology.

While we are yet to understand the causal relation of PARV4 with CNS infection, the findings reveal a high prevalence of the virus, significant co-infection rates with other pathogens, and an association with increased mortality in cases of suspected meningitis. Understanding PARV4’s biology, clinical implications, and potential transmission routes will be essential for informing public health strategies and improving outcomes for vulnerable pediatric populations.

## Data Availability

All data produced in the present study are available upon reasonable request to the authors

## Contributors

DPK and ARM contributed equally to this work. DPK, ARM, NK, and AMT contributed to drafting the first version of the manuscript, data analysis, and figure preparation. NK contributed to analysis, figures, and study design. AMT contributed to study design, genomics analyses, manuscript review and editing. SR, PCD, HR, LT, AR, AAT, DCD, ABJ, MZI, RM, SN, ANUA, and FI contributed to data interpretation, manuscript review, and editing. PCD and AAT contributed to genomic analyses. HR, ABJ, and SN contributed to surveillance activities and study implementation. DCD contributed to laboratory work. MZI, RM, and SN contributed to study implementation and clinical oversight. CT contributed to training and analytical support. JLD, SKS, YH, and SS conceptualised and designed the study and contributed to data interpretation and manuscript revision. SS obtained funding and oversaw project administration and management.

All authors had full access to the data in the study and accept responsibility for the decision to submit for publication.

## Data sharing

The sequence reads of 70 PARV4 genomes are available in the European Nucleotide Archive (ENA) under the study accession PRJEB88738 / ERP171815. All scripts and codes used to create the figures can be found in github folder https://github.com/CHRF-Genomics/PARV4-Analysis

## Acknowledgements

This study was supported by the Gates Foundation and the Child Health Research Foundation, Dhaka, Bangladesh. From the Child Health Research Foundation, we would like to thank Nusrat Alam, and Sharmistha Goswami for helping us with metagenomics and amplicon sequencing and Adittya Arefin for assisting with literature review.

