## appendix for "Neuroinvasive Human Parvovirus 4 is associated with increased mortality in children: a multiyear retrospective observational study"

#### **Text S1. Inclusion criteria for WHO-defined suspected meningitis surveillance**

Children aged 0–59 months admitted to Bangladesh Shishu Hospital and Institute were enrolled in meningitis surveillance according to the World Health Organization (WHO) clinical case definition for suspected meningitis. Suspected meningitis was defined as sudden onset of fever (>100·4°F) accompanied by at least one of the following: neck stiffness; altered or reduced level of consciousness; bulging fontanelle (in children aged <12 months); prostration or lethargy; convulsions; toxic appearance; petechial or purpuric rash; or poor sucking or irritability (in infants aged <2 months). In addition, any child admitted with a clinical diagnosis of meningitis made by the attending physician was eligible for enrolment in the surveillance.

#### **Text S2. Laboratory methods used to process CSF specimens**

Cerebrospinal fluid (CSF) specimens were cultured on chocolate agar, blood agar, and MacConkey agar plates and incubated overnight at 37°C to assess bacterial growth.^1^ For culture-positive specimens, colony morphology was documented and Gram staining was performed. Triple Sugar Iron (TSI), Motility Indole Urea (MIU), and Simmons citrate media were used for presumptive identification of Gram-negative enteric organisms.

CSF infection was assessed by white blood cell (WBC) count using a Neubauer chamber, with differentiation into lymphocytes and polymorphonuclear neutrophils. Protein and glucose concentrations in CSF were measured and recorded. Rapid antigen detection for Streptococcus pneumoniae was performed using an immunochromatographic test (BinaxNOW; Abbott, USA) according to the manufacturer’s instructions.^2,3^ All CSF specimens were subsequently tested for common meningitis-causing pathogens by multiplex quantitative PCR (qPCR), targeting *Neisseria meningitidis* (sodC gene), *Haemophilus influenzae* (hpd gene), and *Streptococcus pneumoniae* (lytA gene).^1^

#### **Text S3. PARV4 and B19 qPCR design and detection**

PARV4 primer design: Primers and probe were designed to target the VP1 region of PARV4. A multiple sequence alignment was constructed using three PARV4 genome sequences generated at CHRF (CHRF_BD0003: ERS29405591, CHRF_RNA_0012: SRR8476518 and CHRF_RNA_0015: SRR8476523) and seven publicly available GenBank sequences: HQ113143·1 and MH215556·1 (both are chimpanzee PARV4 sequences), AY622943·1, DQ873390·1, KM390025·1, KM390024·1 and NC_007018·1 (reference genome). A conserved region between positions 3300–3400 bp (relative to NC_007018·1) within the VP1 gene was identified and used for primer and probe design.

PARV4 detection by qPCR: Quantitative PCR (qPCR) was performed in a 25 µL reaction containing 12·5 µL Perfecta qPCR ToughMix Low ROX (QuantaBio, USA), forward and reverse primers, probe, and 2 µL whole CSF. Amplification was performed on an Applied Biosystems 7500 Fast Dx Real-Time PCR Instrument (Thermo Fisher Scientific, USA). The amplicon size was 95 bp. Primer concentrations were 600 nM each (forward: 5′-CTGATGTTCAACTTTCTCAGGTCCTACCG-3′; reverse: 5′-CCTTCTTGGCTGAGGGCTCTC-3′), and probe concentration was 200 nM (5′-Cy5-CCGCTCCTCCTTCTTCGGACCAACAACCG-BHQ2-3′). Cycling conditions were 50°C for 2 min, 95°C for 10 min, followed by 40 cycles of 95°C for 15 s and 60°C for 1 min.

B19 primer design: A multiple sequence alignment was constructed using four CHRF-derived B19 sequences (CHRF_CSF_0027_TP4: ERS29405606, CHRF_CSF_0020_TP4: ERS29405601, CHRF_CSF_0015_TP4: ERS29405596, CHRF_CSF_0028_TP4: ERS29405607) and 22 GenBank sequences: KM393164·1, KM393163·1, AY504945·1, M24682·1, MN765170·1, FN598217·1, MN765167·1, KT310174·1, KM393168·1, MN765168·1, AF162273·1, KM393166·1, MH201455·1, FJ591158·1, MH201456·1, MT410187·1, KM393165·1, MN765169·1, KM393167·1, KM393169·1, MH151117·1 (reference genome), and AY386330·1. A conserved region between positions 2961–3109 bp (relative to MH151117·1) within the VP1 gene was selected for primer and probe design.

B19 detection by qPCR: Reaction conditions were identical to those described for PARV4. The amplicon size was 86 bp. Primer concentrations were 400 nM each (forward: 5′-TTCAAGGAAGTTTGCCGGA-3′; reverse: 5′-GCTGGCTTCTGTAGAATTAACTGA-3′), and probe concentration was 100 nM (5′-Texas Red-CCCGCTTACAACGCCTCAGAAAAATACCCA-BHQ2-3′). Cycling conditions were 50°C for 2 min, 95°C for 10 min, followed by 40 cycles of 95°C for 15 s and 60°C for 1 min.

#### **Text S4. 16S Sanger sequencing of additional pathogens detected**

Additional organisms detected by culture from CSF were subjected to 16S Sanger sequencing for identification at species level if indefinable through standard microbiological methods. The entire 16S region was covered using forward primer 27F (5’- AGAGTTTGATCMTGGCTCAG-3’) and reverse primer 1492R (5’- CGGTTACCTTGTTACGACTT-3’). For the amplification PCR, in a reaction volume of 25 µl, Hot Firepol 5X master mix (Solis Biodyne, Estonia) was added 5 ul and forward and reverse primers were added at 500 nM final concentration. The PCR cycle was as follows: 15 min at 95℃, 32 cycles of 30 sec at 95℃, 30 sec at 55℃ and 90 sec at 72℃ followed by 5 min at 72℃. Cycle sequencing was done using BigDye terminator v3·1 cycle sequencing kit (Thermo Fisher Scientific, USA) and both forward and reverse primer at final concentration of 500 nM. Cycling conditions were as follows: 1 min at 96, 35 cycles of 10 sec at 96, 5 sec at 50 and 4 min at 60. After bead purification, capillary electrophoresis was done on SeqStudio Flex Genetic Analyzer (Thermo Fisher Scientific, USA). For species detection sequence data was analyzed on BLASTn.

#### **Text S5. Unbiased metagenomic sequencing**

Library preparation was performed using NEBNext Ultra II RNA Library Preparation Kit and the NEBNext Ultra II FS DNA Library Preparation Kit (New England Biolabs, USA) for RNA and DNA metagenomics, respectively. For RNA metagenomics, fragmentation and priming were performed at 94°C for 5 min. Complementary DNA (cDNA) was synthesised by first- and second-strand synthesis, followed by SPRI bead clean-up and size selection at a 1·8× bead ratio. End repair was performed, and hairpin loop-structured adaptors were ligated to the prepared fragments. A 0·9× SPRI bead clean-up step was used to remove unbound adaptors and short fragments. Uracil excision, indexing (barcoding), and PCR enrichment were subsequently performed on adaptor-ligated DNA. Two additional 0·75× SPRI bead clean-up and size-selection steps were undertaken to obtain the final libraries. For DNA metagenomics, enzymatic fragmentation was performed at 37°C for 7 min, followed by library preparation according to the manufacturer’s instructions. Final libraries were sequenced on NextSeq 2000 platform (Illumina, USA) at CHRF, generating 2 × 150 bp paired end reads according to the manufacturer’s protocol.

#### **Text S6. Amplicon sequencing protocol and consensus genome building**

Near-complete PARV4 genomes were amplified using multiplex PCR with primer pools designed by PrimalScheme. Amplification was performed on a Veriti 96-well Fast Thermal Cycler (Applied Biosystems, USA). Each 12·5 µL reaction contained 4·24 µL DNA template, 10 µM of each primer pool, and 6·25 µL NEBNext Ultra II Q5 Master Mix (New England Biolabs, USA). Thermal cycling conditions were 98°C for 30 s, followed by 35 cycles of 98°C for 15 s and 64°C for 5 min, with a final hold at 4°C. Amplicon libraries were prepared using the same protocol described for DNA metagenomics (Text S5; appendix p6). Libraries were sequenced using paired-end 150 bp chemistry on either iSeq 100 or NextSeq 2000 platform (Illumina, USA) according to the manufacturer’s instructions. Consensus genomes were generated from raw FASTQ/FASTA files using the CZ ID viral consensus genome pipeline, an open-source cloud-based bioinformatics platform for next-generation sequencing analysis.^4^

Metagenomic FASTQ files were initially processed using the Illumina DRAGEN Metagenomics pipeline (version 7·1·12) to identify organisms present in each sample. To improve PARV4 genome coverage and ensure a consistent reference across samples, consensus genomes were subsequently generated from metagenomic data by manually exporting FASTA files corresponding to the taxon Tetraparvovirus and importing them into the viral consensus genome pipeline (version 3·4·18). For amplicon-derived data, FASTQ files were directly uploaded to the CZ ID viral consensus genome pipeline (version 3·5·0) together with a Browser Extensible Data (BED) file specifying primer binding positions used for PARV4 enrichment. The consensus genome workflow has been described previously.^4^ Reference-based mapping was performed using GenBank accession KM390024·1, selected as the closest match to study sequences based on BLASTn analysis.

#### **Text S7. Genotyping of PARV4**

A set of 30 reference PARV4 sequences (ten genotype 1, 13 genotype 2, and seven genotype 3) obtained from Matthew and colleagues was used to define genotype-specific signatures*.*^5^ We mapped these sequences against the reference genome (GenBank accession: NC007018·1.) using minimap2 to identify Single Nucleotide Polymorphisms (SNPs).^6^ Variant Call Format (VCF) files were generated through SNP analysis as described earlier.^7^

All detected SNPs were tracked to identify genotype-specific SNP signatures for genotype-1, 2, and 3, within conserved protein-coding sequences. The defining SNPs for genotype 1 were located at the nucleotide position 700, 1076, 1129, and 5119 with bases A, G, A, A; for genotype 2, at 3241, 3434, 3526, 3574, 3592, and 3652 with bases G, A, G, T, A, A; and for genotype 3, at 2623, 2647, 2690, 2749, 2794, 3154, 3232, 3259, and 3286 with bases G, C, T, T, T, T, T, T, T, C. We used these unique SNP signatures to develop a customized genotyping tool, written in Python (named Tetratype; <https://github.com/CHRF-Genomics/PARV4-Analysis.git>) to determine genotypes of our PARV4 sequences.

#### **Text S8. Estimating genome-wide substitutions/year**

We used the whole-genome alignment with MAFFT and BEAST v1·10·4^8^ to perform Bayesian phylogenetic analysis under the lognormal uncorrelated relaxed clock, varying substitution-model parameters and tree priors (Table S5). Independent MCMC chains were run for 100 or 200 million steps each, sampling every 50 000 iterations. Resulting log files were assessed in Tracer v1·7·1, and the “joint”, “ucldMean” (reported as clock.rate in the Table), and “treeLength” values were extracted with their 95 % highest posterior density (HPD) intervals and effective sample sizes (ESS). The “joint” values were extracted to evaluate the overall quality of each MCMC run. Here, the “clock.rate” refers to the average substitution rate per site per year across the alignment. Next, we converted this per-site rate to genome-wide substitutions per year using the formula below:

$$Genomewide substitutions per year=\frac{(Posterior mean of clock.rate) \times(Alignment length in bp)}{Number of Year}$$

The alignment length was 5,284. We selected the genome-wide substitutions per year estimates from the MCMC run with the lowest “joint” value.

#### **Text S9. Mutation profiling**

Two open reading frames (ORF(s)) of PARV4 expressing NS1 (ORF1) and VP1 and VP2 (ORF2) were analyzed for non-synonymous (NS) mutations. All our sequences were processed using Biopython v1·8 and translated using Prokka v1·14·6 to analyze them at amino-acid level. Samples containing long stretches of unidentified nucleotides in either of the ORFs, which prevented successful translation, were excluded that resulted in a total of 67 samples for downstream comparative analysis.

CHRF_CSF_0090 (accession number: ERS29405649) was used as the reference genome when defining NS mutations and subjected to tBLASTn on NCBI against [AY622943](https://www.ncbi.nlm.nih.gov/nuccore/AY622943) to identify ORF positions and annotate coding regions (NS1, VP1 and VP2).^9^ Additionally, previously defined variable regions (VR-I to VR-IX and the HI loop) were treated as VP2 based amino acid coordinates. NS mutation was recorded when an amino acid in the sample differed from the amino acid in the reference sequence at a given position. A missing amino acid in either of the sequences was ignored, and any ambiguous amino acid was masked during visualization to prevent misinterpretation of mutation.

For each ORF, a mutation matrix was constructed, and the proportion of NS mutations at each site of the genome was calculated relative to the reference genome using NumPy v1·26·4. At each amino acid position, the proportion of NS mutation was defined as:

$Proportion of NS mutation= \frac{Number of sample with NS mutation}{Number of valid sequences at that position}x 100$

All sequences with an amino acid at a given position were considered valid. Plots of mutation profiling were visualized using Matplotlib v3·9·2.

### Supplementary Figures

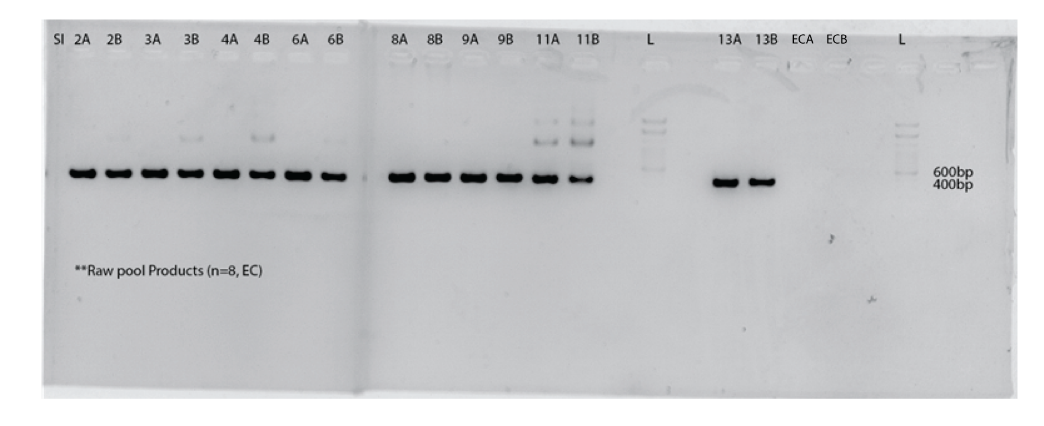

#### **Figure S1. Primer pool validation of PARV4**

Eight CSF samples positive for PARV4 by qPCR with Ct <25 were selected for validation. DNA was extracted using QIAamp DNA mini kit (Qiagen, Germany) following quality check with Nanodrop One (Thermo Fisher Scientific, USA). PARV4 pool A and pool B primers were diluted to 10 µM. Multiplex PCR was performed following the ARTIC-NEB: SARS-CoV-2 Library prep V.4 (<https://www.protocols.io/view/artic-sars-cov-2-sequencing-protocol-v4-lsk114-bp2l6n26rgqe/v4>). Briefly, 4·25 µl of DNA was mixed with 6·25 µl of NEBNext Ultra II Q5 master mix (New England Biolabs, USA) in a reaction volume of 12·5 µl. From each pool, 10 µM primer was used. PCR was done on the Veriti 96-well Fast Thermal Cycler (Applied Biosystems) and the cycling conditions were as follows: 30 sec at 98°C; 30 cycles of 15 sec at 98°C and 5 min at 64°C, followed by hold at 4°C. Multiplex pool PCR products were run in 1·5% agarose gel at 100V for 50 min. Distinct bands were observed at 400 bp. L = 1kb DNA ladder (Invitrogen, USA) . EC= extraction control

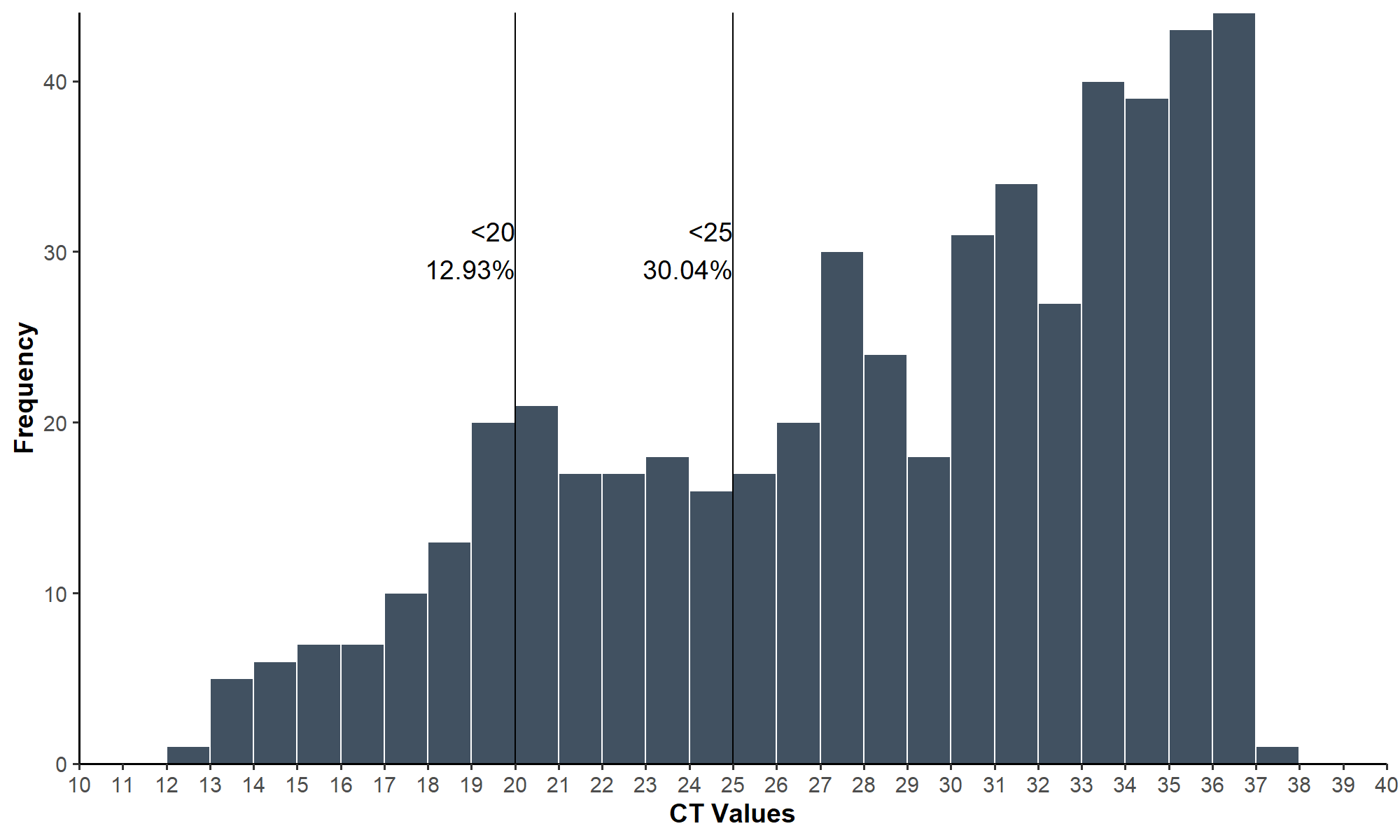

#### **Figure S2. Distribution of cycle threshold (Ct) values among PARV4-positive cases**

Histogram showing the distribution of Ct values. Vertical lines indicate thresholds of Ct <20 and Ct <25, with corresponding proportions shown.

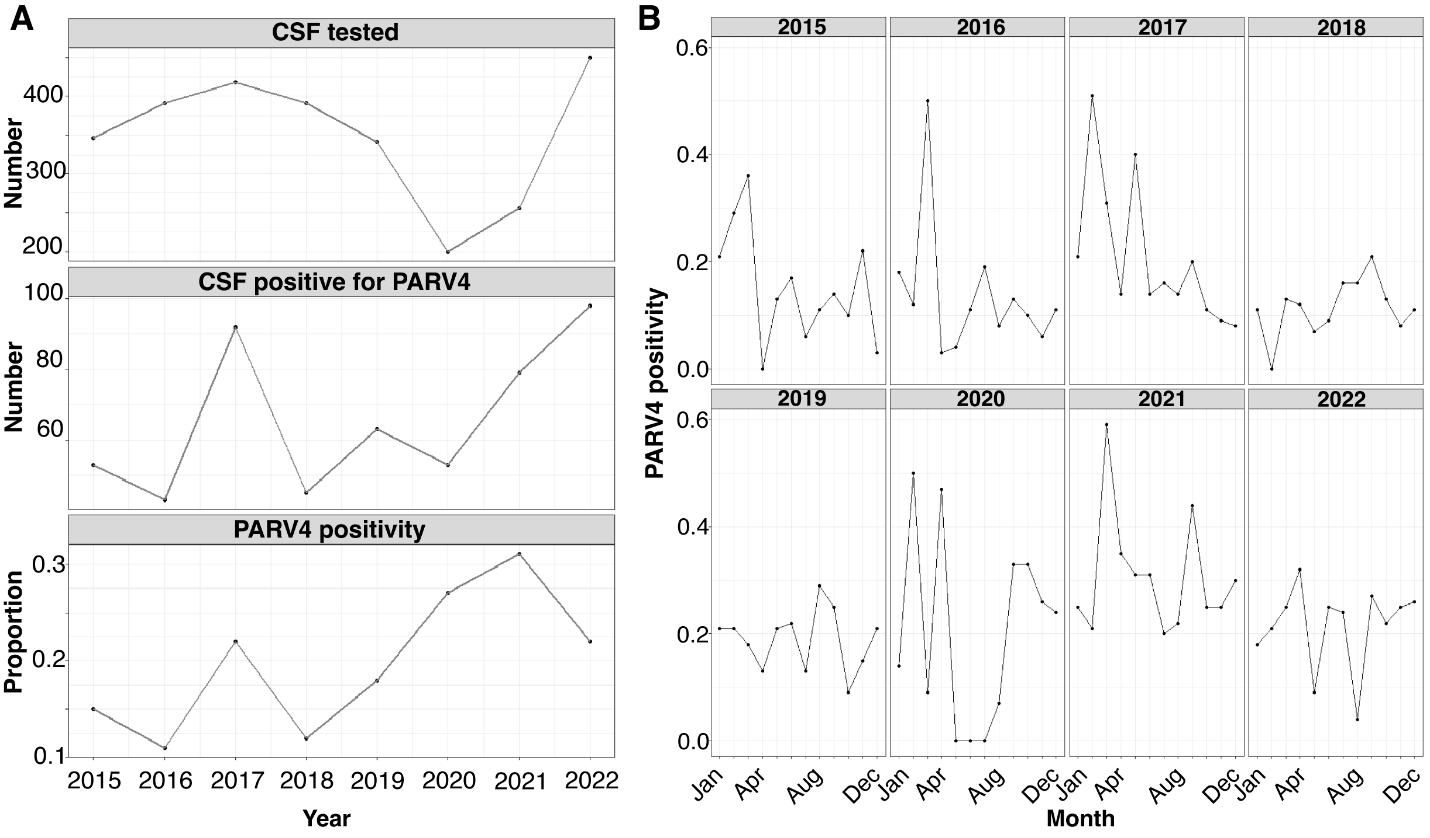

#### **Figure S3. Prevalence of PARV4 positivity in suspected meningitis cases**

**(A)** Annual prevalence of PARV4 positivity from 2015 to 2022. Highest positivity was seen in 2021 (31%) and lowest positivity was seen in 2016 (11%). **(B)** Monthly distribution of PARV4 positivity across the study period.

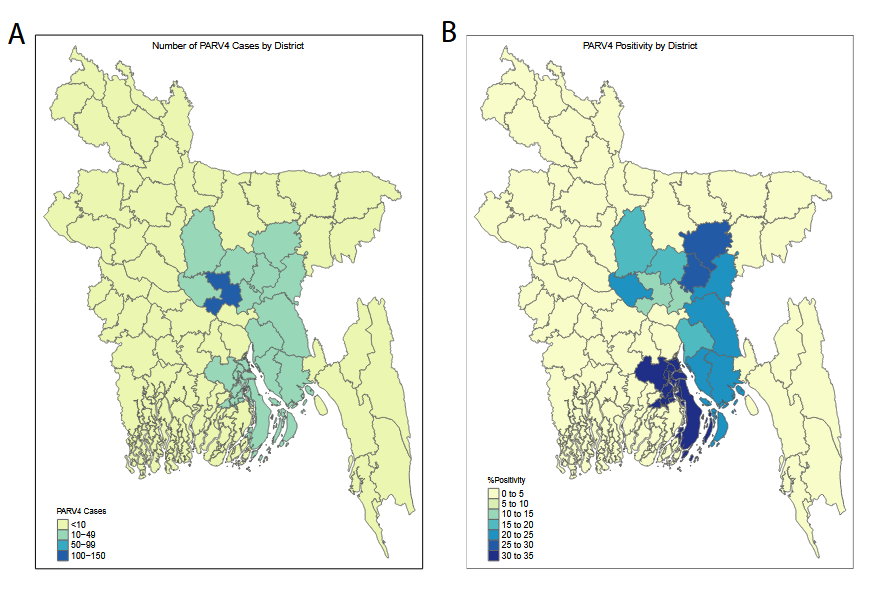

#### **Figure S4. Geographic distribution of PARV4 positivity and case counts by district**

**(A)** Number of PARV4-positive cases by district. This map displays the absolute number of PARV4-positive cases reported per district, with color intensity corresponding to case count categories: less than 10, 10–49, 50–99, and 100–150 cases. **(B)** Percentage of PARV4 positivity by district. This map shows the distribution of PARV4 positivity rates across districts in Bangladesh. Darker shades represent higher positivity percentages, with rates ranging from 0% to over 30%, indicating geographic variability in PARV4 prevalence.

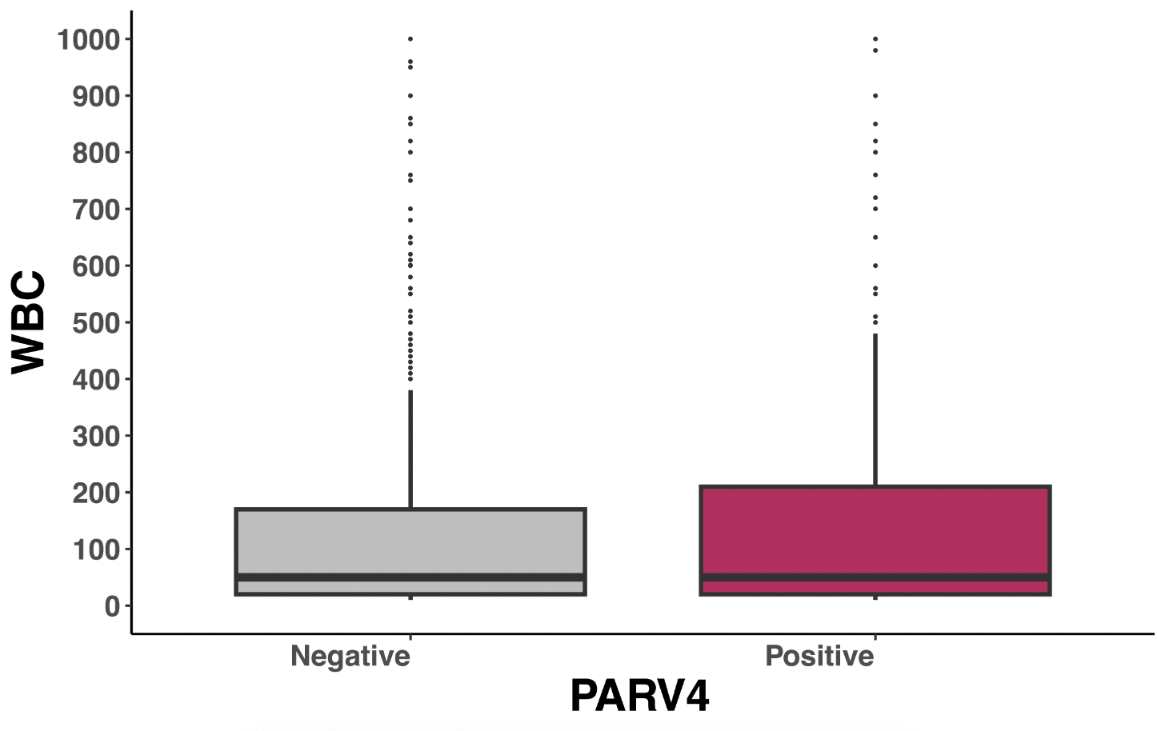

#### **Figure S5. White Blood Cell (WBC) count in PARV4-positive and PARV4-negative suspected meningitis cases**

This box plot compares the distribution of WBC counts in CSF samples from PARV4-positive and PARV4-negative pediatric meningitis cases. The median WBC count is indicated by the central line within each box, with the IQR represented by the box edges. Whiskers extend to 1·5 times the IQR, and dots represent outliers. PARV4-positive cases show a similar WBC distribution to PARV4-negative cases, suggesting no significant difference in WBC counts between the groups.

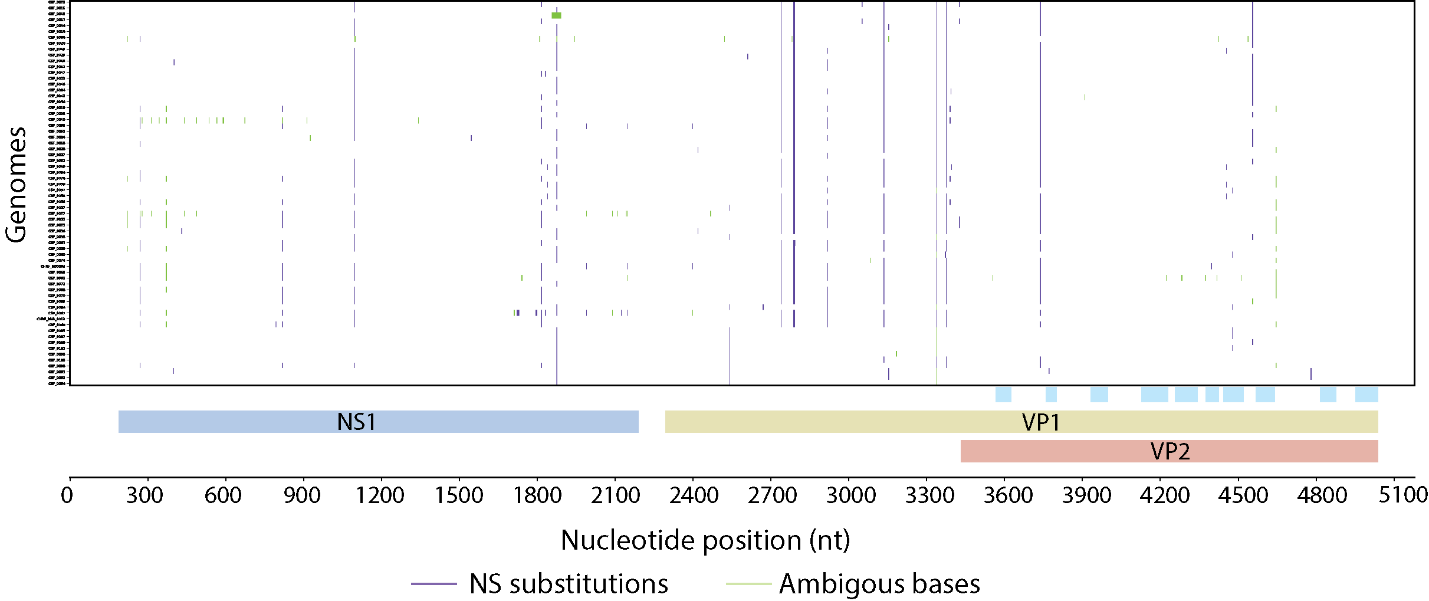

#### **Figure S6. Genome-wide distribution of non-synonymous variation across PARV4 genomes**

Genome-wide non-synonymous (NS) variation across 66 PARV4 genomes with complete ORF1 and ORF2 sequences. The x-axis indicates genome position (nt) corresponding to NS1 (aa 63–730; nt 190–2190), VP1 (aa 765–1680; nt 2296–5037), and VP2 (aa 1127–1680; nt 3382–5037). Each row represents an individual genome. Vertical coloured lines denote sequence variation: purple, amino acid substitutions; green, ambiguous amino acids. Variable regions within VP2 (VR-I–VR-IX and the HI loop) are indicated by blue bars.

### Supplementary tables

#### **Table S1. Design of PARV4 primer pool**

Initially, we designed two sets of pool primers using Primal Scheme based on the alignment of either our sequences from unbiased metagenomics or global sequences from NCBI. Seven sequences obtained from unbiased metagenomics were aligned (CHRF_CSF_0017_TP4, CHRF_CSF_0004, CHRF_CSF_0018_TP4, CHRF_CSF_0019_TP4, CHRF_CSF_0023_TP4, CHRF_CSF_0026_TP4, and including the reference CHRF_BD0003) that generated a set of sixteen pool 1 primers and fourteen pool 2 primers. However, 4912 bp long reference genome was only partially covered by these pool primers resulting in incomplete coverage at both ends. To fill in these gaps we generated four additional pool 1 primers from the alignment of 5 global sequences from NCBI (KM390024·1, AY622943·1, EU874248·1, HQ593530·1, and including the reference NC_007018·1). The revised set of pool primers consisted of twenty pool 1 primers and fourteen pool 2 primers with the potential to amplify the complete genome of PARV4. All alignments were generated using Clustal Omega.^10^ Both Primal Scheme runs aimed to generate 400 bp long amplicons. Accession number of our sequences from unbiased metagenomics: ERS29405598 (CHRF_CSF_0017_TP4), ERS29405595

(CHRF_CSF_0004), ERS29405599 (CHRF_CSF_0018_TP4), ERS29405600 (CHRF_CSF_0019_TP4), ERS29405603 (CHRF_CSF_0023_TP4), ERS29405605 (CHRF_CSF_0026_TP4), ERS29405591 (CHRF_BD0003).

| **Original Positions** | | | **Corresponding positions on NC_007018·1** | | | **Pool** | **Strand** | **Complete match to Ref (Yes/No)** | **Sequence 5' to 3'** | **Length (bp)** |
| --- | --- | --- | --- | --- | --- | --- | --- | --- | --- | --- |
| **Ref** | **start** | **end** | **Ref** | **start** | **end** |  |  |  |  |  |
| NC_007018·1 | 85 | 105 | NC_007018·1 | 85 | 105 | 1 | + | Yes | TTTCCGGCCACGTCAACTTC | 20 |
| NC_007018·1 | 463 | 485 | NC_007018·1 | 463 | 485 | 1 | - | Yes | GCAAACAGCTGTTGCAATATGC | 22 |
| CHRF_BD0003 | 62 | 83 | NC_007018·1 | 307 | 327 | 1 | + | Yes | GTGCTGCAAATTCCTACCGGA | 21 |
| CHRF_BD0003 | 437 | 464 | NC_007018·1 | 682 | 708 | 1 | - | Yes | ACGGGTTTTTCTAATATTCCAGCTTAA | 27 |
| CHRF_BD0003 | 361 | 383 | NC_007018·1 | 606 | 627 | 2 | + | Yes | AGGCAGAGAATGTAGCAACTGG | 22 |
| CHRF_BD0003 | 733 | 758 | NC_007018·1 | 978 | 1002 | 2 | - | Yes | TTCTGTGGCTATTCCATTTTCAACT | 25 |
| CHRF_BD0003 | 669 | 691 | NC_007018·1 | 914 | 935 | 1 | + | Yes | AAGGAGTTCCCCCTACTATGGC | 22 |
| CHRF_BD0003 | 1051 | 1073 | NC_007018·1 | 1300 | 1317 | 1 | - | No | AGCAGCTAACAGAGTTTTGCCA | 22 |
| CHRF_BD0003 | 976 | 997 | NC_007018·1 | 1221 | 1241 | 2 | + | Yes | ATGGTATTTTGCTGCATGGGC | 21 |
| CHRF_BD0003 | 1359 | 1384 | NC_007018·1 | 1604 | 1628 | 2 | - | Yes | TGAAACATGGTTATTCTTGTCTGCA | 25 |
| CHRF_BD0003 | 1290 | 1315 | NC_007018·1 | 1537 | 1551 | 1 | + | No | TCACCTCTAATGGAGATCTTACGGT | 25 |
| CHRF_BD0003 | 1680 | 1703 | NC_007018·1 | 1925 | 1943 | 1 | - | No | ACTTGGAGAGGAGAGCTCAATGA | 23 |
| CHRF_BD0003 | 1616 | 1638 | NC_007018·1 | 1861 | 1882 | 2 | + | Yes | CCACCACCCACTCAGAAGAAAA | 22 |
| CHRF_BD0003 | 1996 | 2025 | NC_007018·1 | 2245 | 2269 | 2 | - | No | ACCAAGCCCAAATAAATCTTCTATTAACC | 29 |
| CHRF_BD0003 | 1938 | 1960 | NC_007018·1 | 2185 | 2204 | 1 | + | No | TGTGGTCTCGAGAGTTTTGGAG | 22 |
| CHRF_BD0003 | 2303 | 2326 | NC_007018·1 | 2547 | 2569 | 1 | - | Yes | AGCAAATTTAAGCAAGCCTTGGA | 23 |
| CHRF_BD0003 | 2224 | 2246 | NC_007018·1 | 2468 | 2489 | 2 | + | Yes | GAACCTTCAAGTCCAGCGAACC | 22 |
| CHRF_BD0003 | 2607 | 2630 | NC_007018·1 | 2851 | 2862 | 2 | - | No | TGCCAAAATTTGTTAGCGAGTCT | 23 |
| CHRF_BD0003 | 2539 | 2559 | NC_007018·1 | 2784 | 2796 | 1 | + | No | CGAGGCATTGTGCGATTTGC | 20 |
| CHRF_BD0003 | 2916 | 2938 | NC_007018·1 | 3160 | 3181 | 1 | - | Yes | AGCGCCGTGACCATGTAAATAA | 22 |
| CHRF_BD0003 | 2850 | 2872 | NC_007018·1 | 3094 | 3115 | 2 | + | Yes | AGTGGATGAGGCAGCAAAACAT | 22 |
| CHRF_BD0003 | 3236 | 3258 | NC_007018·1 | 3480 | 3501 | 2 | - | Yes | TTAACTCCACCTCCTCCTCCAG | 22 |
| CHRF_BD0003 | 3163 | 3185 | NC_007018·1 | 3407 | 3428 | 1 | + | Yes | ATTGGTACCCCTGAGGAGTCTG | 22 |
| CHRF_BD0003 | 3556 | 3578 | NC_007018·1 | 2727 | 2737 | 1 | - | No | TGGTGCCTGTCTGATTAGTTGC | 22 |
| CHRF_BD0003 | 3460 | 3488 | NC_007018·1 | 979 | 987 | 2 | + | No | ACTCCCAGAGATTTTCAACAATTACTAG | 28 |
| CHRF_BD0003 | 3854 | 3879 | NC_007018·1 | 4098 | 4122 | 2 | - | Yes | TTTCTCCAAGGTAGATCATCTGGAA | 25 |
| CHRF_BD0003 | 3795 | 3817 | NC_007018·1 | 4043 | 4060 | 1 | + | No | AGAACACCATGATGCAGAGTGT | 22 |
| CHRF_BD0003 | 4164 | 4186 | NC_007018·1 | 4412 | 4429 | 1 | - | No | TGCTAACCATTCTTCTGTGCGA | 22 |
| CHRF_BD0003 | 4090 | 4113 | NC_007018·1 | 4334 | 4356 | 2 | + | Yes | CCTGTTGCTATTGGAAATCCTGC | 23 |
| CHRF_BD0003 | 4474 | 4504 | NC_007018·1 | 4718 | 4747 | 2 | - | Yes | AGTCCATATTTGACAGTCATATGATAAAGG | 30 |
| CHRF_BD0003 | 4412 | 4437 | NC_007018·1 | 4656 | 4680 | 1 | + | Yes | GCCCAAAATTGTATCAAGAACCTGT | 25 |
| CHRF_BD0003 | 4795 | 4819 | NC_007018·1 | 5039 | 5055 | 1 | - | No | CGTGTTTGGGTCTTGTAAGCTATT | 24 |
| NC_007018·1 | 4705 | 4734 | NC_007018·1 | 4705 | 4734 | 1 | + | Yes | TGGAACCCAAATCCTTTATCATATGATTG | 29 |
| NC_007018·1 | 5086 | 5108 | NC_007018·1 | 5086 | 5108 | 1 | - | Yes | AGTAATTGCGCGCAATCCATTG | 22 |

#### **Table S2. Time from admission to sample collection**

Time of sample collection was available for 2792 of 2793 samples.

| **The interval between time of admission and sample collection** | **Number of Samples** | **Percentage** |
| --- | --- | --- |
| 0 - 23 hours | 834 | 29·87 |
| 24 - 47 hours | 603 | 21·60 |
| 48 - 71 hours | 362 | 12·97 |
| >=72 hours | 993 | 35·57 |

#### **Table S3. Assigned clusters of related diagnoses**

Respiratory manifestation

Pneumonia/ bronchopneumonia

Severe pneumonia

Bronchiolitis/ acute bronchiolitis

ARI/Acute respiratory infection

RDS/ Respiratory distress syndrome

Systemic infections

Septicaemia / sepsis

Neonatal sepsis

Febrile illness

Enteric fever/ typhoid fever / paratyphoid fever

Febrile convulsion / atypical febrile convulsion

Viral fever/dengue Fever

Neurological manifestation

Meningitis

Encephalitis/meningoencephalitis/encephalopathy

Seizure disorder/neonatal seizure

Epilepsy

Gastrointestinal manifestation

Acute gastroenteritis/ AGE/acute watery diarrhea

Persistent diarrhoea

Dysentery/ shigellosis/ invasive diarrhoea

Genitourinary/renal manifestation

Nephrotic syndrome

Renal failure/ kidney failure

AGN/ acute glomerulonephritis/ APSGN

Cardiovascular manifestation

Ventricular septal defect/ VSD

Tetralogy of fallot/ TOF

Other congenital heart disease

Perinatal asphyxia

Preterm low-birth weight

Neonatal jaundice

Other*

*****Other may include the following diagnoses:

| Neonatal jaundice |  | Club foot/ TEV |
| --- | --- | --- |
| Neurological manifestation |  | CMV infection |
| Genitourinary/renal manifestation |  | Collodion baby |
| Congenital Cardiovascular manifestation |  | Congenital anomaly/ cloacal anomaly |
| Acute abdomen |  | Congenital pneumonia |
| ABO incompatibility/ Rh incompatibility |  | Congenital rubella syndrome |
| Achondroplasia |  | Congenital syphilis |
| Acute Appendicitis/ burst appendix |  | Constipation |
| Acute Epididymoorchitis/ orchitis |  | Cough & cold/common cold |
| Acute flaccid paralysis/ AFP |  | Croup |
| Acute gastritis |  | Cystic fibrosis |
| Acute laryngotracheobronchitis |  | Cystic hygroma |
| Acute leukemia (ALL/AML) |  | Dengue fever |
| Acute otitis media/otitis media (ASOM) |  | Dermatitis |
| Acute scrotum |  | Developmental delay |
| Acute Severe Asthma/acute exacerbation |  | Diabetes/ diabetes mellitus |
| Acute stroke syndrome |  | Down's syndrome |
| Adrenal Hyperplasia |  | Drug overdose/drug reaction/poisoning |
| AEFI/ adverse effect following immunization |  | Electrolyte imbalance |
| Anaemia |  | Empyema thoracis |
| Anal fissure |  | Encephalopathy |
| Anorectal malformation |  | Eventration of diaphragm |
| Anuria/ oliguria |  | Extrapulmonary Tuberculosis |
| Aplastic anemia |  | Failure to thrive |
| Arthritis/ rheumatoid arthritis/ JIA |  | Feeding problem/feeding mismanagement |
| Ascariasis/ helminthiasis |  | Fistula |
| Aspiration Pneumonia |  | G-6-PD Deficiency |
| Aspiration syndrome |  | Gastro esophageal reflux disease/ GERD |
| Ataxia/ cerebellar ataxia |  | Growing pain |
| Bell's palsy/ facial nerve palsy |  | Guillain- Barre syndrome/GBS |
| Biliary atresia |  | Gut atresia/ choanal atresia/ intestinal |
| Birth injury |  | Gut malrotation |
| Bladder Obstruction/ urinary obstruction |  | Haemorrhagic diseases of new born (HDN) |
| Bleeding disorder |  | HB-E Diseases |
| Breath holding attack |  | Head injury |
| bullous disorder/ Epidermolysis Bullosa |  | Healthy baby/ Normal baby/ well baby |
| Burn/ scald/ post burn contracture |  | Heart failure |
| Cardiomyopathy/ cardiomegaly |  | Hemangioma |
| Cellulitis |  | Hematemesis |
| Cerebral atrophy/ cortical atrophy |  | Hematoma/ cephalohematoma |
| Cerebral malaria/malaria/severe malaria |  | Hemiplegia |
| Cerebral palsy |  | Hemolytic anemia |
| Chicken pox |  | Hepatitis/neonatal hepatitis Syndrome |
| Chocking/ chocking Attack |  | Hepatoblastoma |
| Cholecystitis/ choledochal cyst |  | Hernia/ Inguinal hernia |
| Chronic kidney disease/ CKD |  | Hypoxic-ischaemic encephalopathy |
| Chronic suppurative otitis media (CSOM) |  | Hirschsprung disease/ HD |
| CLD/chronic liver disease |  | Hypertension |
| Cleft lip/Cleft palate |  | Hydrocele/ Encysted hydrocele |
| Hydrocephalus |  | Per rectal bleeding |
| Hydronephrosis |  | Pericardial effusion |
| Hypoglycemia |  | Pharyngitis |
| Hypospadias |  | Phimosis/ para phimosis |
| Hypothyroidism |  | Pierre robin syndrome |
| ICSOL/ Intracranial space occupying lesions |  | Pleural effusion |
| Ichthyosis |  | Pneumonic consolidation |
| IDM/ infant of diabetic mother |  | Pneumonitis |
| Infantile colic/ infantile spasm |  | Pneumothorax/Hydro pneumothorax |
| Infantile hypertrophic pyloric stenosis |  | Polycystic kidney disease/ PKD |
| Infected scabies/scabies |  | Portal HTN/ portal hypertension |
| Infective endocarditis |  | Post circumcision bleeding |
| Injury/ Cut injury/ lacerated injury |  | Post measles pneumonia |
| Insect bite/ bee bite |  | Post meningitis neurological sequalae |
| Intestinal obstruction/ paralytic ileus |  | Postdated baby |
| Intestinal perforation |  | Pseudotumor cerebri |
| Intestinal tuberculosis |  | Pulmonary hypertension |
| Intrauterine Growth retardation (IUGR) |  | Pure red cell aplasia |
| Intussusception |  | PUV/ posterior urethral valve |
| ITP /Idiopathic thrombocytopenic Purpura |  | Pyrexia of unknown origin (PUO) |
| Jaundice |  | Rabies |
| Jejunal atresia |  | Reactive airway disease |
| Kala azar |  | Rectal polyp |
| Laryngomalacia |  | Respiratory wheeze/recurrent wheeze |
| Lipoma |  | Rheumatic fever |
| Liver abscess/ amoebic liver abscess |  | Rickets |
| Liver failure/ hepatic failure |  | Rickettsial fever |
| Low Birth weight (LBW) |  | Scarlet fever |
| Lung abscess |  | Septic arthritis |
| Lung collapse |  | Sequelae |
| Lymphadenitis/ lymphadenopathy |  | Some/severe dehydration |
| Lymphoma/ Hodgkin's disease |  | Stevens Johnson Syndrome |
| Measles |  | Stomatitis/ Gingivostomatitis |
| Meconium aspiration syndrome (MAS) |  | Storage disease |
| Medulloblastoma |  | Sub dural effusion |
| Malingering |  | Subacute sclerosing panencephalitis |
| Meningocele/ myelocele/ meningomyelitis |  | Synovitis/ transient synovitis |
| Meningoencephalitis |  | Teratoma |
| Microcephaly |  | Transposition of great artery |
| Motor neuron disease/ UMN |  | Thalassemia |
| Mumps |  | Tonsilitis |
| Muscular atrophy/ muscular dystrophy |  | TORCH infection |
| Myopathy/ Myositis/ myalgia |  | Transient tachypnea of newborn |
| Near drowning |  | Tubercular meningitis |
| Neonatal convulsion/neonatal seizure |  | Tuberculosis/disseminated tuberculosis |
| Nephroblastoma |  | Umbilical Hernia |
| Neuroblastoma |  | Umbilical sepsis |
| Neurodegenerative disorder |  | Undescendent testis |
| Neurometabolic disease |  | Upper respiratory tract infection |
| Omphalitis |  | Urinary tract infection |
| Omphalocele |  | Urine suppression |
| Oral Thrush |  | Urticaria/ allergic reaction |
| Osteomyelitis/ acute Osteomyelitis |  | Valvular heart disease |
| Pancreatitis/ acute pancreatitis/ chron |  | Whopping cough |
| PEM/2° PEM |  | Wilms tumor |
| Peptic ulcer disease/ PUD |  |  |

#### **Table S4. Overview of consensus genomes from amplicon sequencing**

| **Sample Name** | **Accession** | **Reference Accession ID** | **Reference Length** | **% Genome Called** | **Total Reads** | **Mapped Reads** | **Coverage Depth** |
| --- | --- | --- | --- | --- | --- | --- | --- |
| CHRF_CSF_0044_TP4 | ERS29405616 | KM390024·1 | 5268 | 98·7 | 305200 | 272286 | 7121·2x |
| CHRF_CSF_0045_TP4 | ERS29405617 | KM390024·1 | 5268 | 99·1 | 463488 | 425572 | 10380·3x |
| CHRF_CSF_0046_TP4 | ERS29405618 | KM390024·1 | 5268 | 99·1 | 302152 | 276328 | 6710·2x |
| CHRF_CSF_0047_TP4 | ERS29405619 | KM390024·1 | 5268 | 99·2 | 497826 | 468731 | 11711·6x |
| CHRF_CSF_0048_TP4 | ERS29405620 | KM390024·1 | 5268 | 98·7 | 371718 | 277038 | 6724·2x |
| CHRF_CSF_0049_TP4 | ERS29405621 | KM390024·1 | 5268 | 98·7 | 220596 | 209856 | 5305·1x |
| CHRF_CSF_0050_TP4 | ERS29405622 | KM390024·1 | 5268 | 98·7 | 195536 | 158699 | 4076·8x |
| CHRF_CSF_0051_TP4 | ERS29405623 | KM390024·1 | 5268 | 99·1 | 591238 | 576003 | 15263·7x |
| CHRF_CSF_0052_TP4 | ERS29405624 | KM390024·1 | 5268 | 99·4 | 116913 | 856054 | 17350·2x |
| CHRF_CSF_0053_TP4 | ERS29405625 | KM390024·1 | 5268 | 99·2 | 803990 | 547065 | 13961·8x |
| CHRF_CSF_0054_TP4 | ERS29405626 | KM390024·1 | 5268 | 99·3 | 661848 | 612425 | 15223·1x |
| CHRF_CSF_0055_TP4 | ERS29405627 | KM390024·1 | 5268 | 99·1 | 628432 | 591388 | 14738·0x |
| CHRF_CSF_0056_TP4 | ERS29405628 | KM390024·1 | 5268 | 99·1 | 571820 | 547718 | 14277·4x |
| CHRF_CSF_0057_TP4 | ERS29405629 | KM390024·1 | 5268 | 99·2 | 625612 | 608518 | 16224·6x |
| CHRF_CSF_0058_TP4 | ERS29405630 | KM390024·1 | 5268 | 99·8 | 706358 | 677498 | 17779·9x |
| CHRF_CSF_0059_TP4 | ERS29405631 | KM390024·1 | 5268 | 99·2 | 528464 | 512571 | 13756·2x |
| CHRF_CSF_0060_TP4 | ERS29405632 | KM390024·1 | 5268 | 98·8 | 165235 | 1207929 | 24857·5x |
| CHRF_CSF_0061_TP4 | ERS29405633 | KM390024·1 | 5268 | 99·2 | 825166 | 721279 | 16966·7x |
| CHRF_CSF_0062_TP4 | ERS29405634 | KM390024·1 | 5268 | 99·1 | 971406 | 891917 | 21718·0x |
| CHRF_CSF_0063_TP4 | ERS29405635 | KM390024·1 | 5268 | 99·8 | 184405 | 1499324 | 31248·9x |
| CHRF_CSF_0064_TP4 | ERS29405636 | KM390024·1 | 5268 | 99·2 | 104514 | 902331 | 20741·3x |
| CHRF_CSF_0065_TP4 | ERS29405637 | KM390024·1 | 5268 | 99·2 | 110456 | 1001409 | 23640·6x |
| CHRF_CSF_0066_TP4 | ERS29405638 | KM390024·1 | 5268 | 98·6 | 912232 | 831552 | 20423·8x |
| CHRF_CSF_0067_TP4 | ERS29405639 | KM390024·1 | 5268 | 99·2 | 118434 | 1033775 | 23532·9x |
| CHRF_CSF_0068_TP4 | ERS29405640 | KM390024·1 | 5268 | 99·8 | 106162 | 927476 | 22056·3x |
| CHRF_CSF_0069_TP4 | ERS29405641 | KM390024·1 | 5268 | 98·7 | 111971 | 968996 | 23461·3x |
| CHRF_CSF_0070_TP4 | ERS29405642 | KM390024·1 | 5268 | 99·2 | 140775 | 1268826 | 29078·0x |
| CHRF_CSF_0071_TP4 | ERS29405643 | KM390024·1 | 5268 | 99·5 | 101071 | 822237 | 18644·8x |
| CHRF_CSF_0072_TP4 | ERS29405644 | KM390024·1 | 5268 | 99·7 | 622988 | 526650 | 12832·1x |
| CHRF_CSF_0074_TP4 | ERS29405645 | KM390024·1 | 5268 | 99·5 | 775042 | 699133 | 18090·2x |
| CHRF_CSF_0075_TP4 | ERS29405646 | KM390024·1 | 5268 | 99·2 | 117200 | 915115 | 19291·4x |
| CHRF_CSF_0076_TP4 | ERS29405647 | KM390024·1 | 5268 | 99·9 | 964950 | 818481 | 17838·2x |
| CHRF_CSF_0077_TP4 | ERS29405648 | KM390024·1 | 5268 | 98·9 | 976198 | 684599 | 14985·3x |
| CHRF_CSF_0036_TP4 | ERS29405608 | KM390024·1 | 5268 | 98·7 | 218200 | 209140 | 5490·2x |
| CHRF_CSF_0037_TP4 | ERS29405609 | KM390024·1 | 5268 | 99·2 | 574708 | 542402 | 13803·2x |
| CHRF_CSF_0038_TP4 | ERS29405610 | KM390024·1 | 5268 | 98·5 | 519344 | 486178 | 12482·7x |
| CHRF_CSF_0039_TP4 | ERS29405611 | KM390024·1 | 5268 | 99·1 | 398402 | 375801 | 9696·1x |
| CHRF_CSF_0040_TP4 | ERS29405612 | KM390024·1 | 5268 | 98·1 | 254872 | 233704 | 5974·1x |
| CHRF_CSF_0041_TP4 | ERS29405613 | KM390024·1 | 5268 | 99·2 | 436218 | 382445 | 9256·2x |
| CHRF_CSF_0042_TP4 | ERS29405614 | KM390024·1 | 5268 | 99·9 | 838188 | 663509 | 15497·1x |
| CHRF_CSF_0043_TP4 | ERS29405615 | KM390024·1 | 5268 | 98·5 | 463226 | 415157 | 10150·0x |
| CHRF_CSF-0090-TP4 | ERS29405649 | KM390024·1 | 5268 | 98·3 | 174142 | 150414 | 3365·6x |
| CHRF_CSF-0091-TP4 | ERS29405650 | KM390024·1 | 5268 | 98·8 | 172542 | 144177 | 3123·1x |
| CHRF_CSF-0092-TP4 | ERS29405651 | KM390024·1 | 5268 | 98·7 | 202012 | 171691 | 3711·3x |
| CHRF_CSF-0093-TP4 | ERS29405652 | KM390024·1 | 5268 | 90·2 | 141490 | 72307 | 1694·0x |
| CHRF_CSF-0094-TP4 | ERS29405653 | KM390024·1 | 5268 | 99 | 274218 | 236248 | 5253·2x |
| CHRF_CSF-0095-TP4 | ERS29405654 | KM390024·1 | 5268 | 98·5 | 213576 | 133873 | 2948·2x |
| CHRF_CSF_0096_TP4 | ERS29405655 | KM390024·1 | 5268 | 99 | 151774 | 136944 | 3245·3x |
| CHRF_CSF_0097_TP4 | ERS29405656 | KM390024·1 | 5268 | 98·6 | 227218 | 200723 | 4444·0x |
| CHRF_CSF_0098_TP4 | ERS29405657 | KM390024·1 | 5268 | 97·7 | 168930 | 150205 | 3496·5x |
| CHRF_CSF_0099_TP4 | ERS29405658 | KM390024·1 | 5268 | 98·5 | 172994 | 159067 | 3707·8x |
| CHRF_CSF_0100_TP4 | ERS29405659 | KM390024·1 | 5268 | 98·3 | 180050 | 157205 | 3635·0x |
| CHRF_CSF_0101_TP4 | ERS29405660 | KM390024·1 | 5268 | 97·8 | 175850 | 150918 | 3352·5x |

#### **Table S5. BEAST MCMC run metrics (joint values, ucldMean (clock.rate), and treeLength estimates with 95 % HPD intervals) and genome wide substitutions/year for each independent chain.**

The following parameters were used for the tree priors and substitution-models; Bayesian Skygrid (number of parameters = 50, time at last transition point = 25·0), Bayesian Skyline (default settings), HKY and GTR (base frequency = empirical, site heterogeneity model = G4+I). The tree prior with the lowest “joint” value (marked with *) was selected for calculation of genome-wide substitutions per year.

| **Tree prior** | **Substitution model** | **MCMC chain length (millions)** | **joint (95% HPD)** | **treelength (95% HPD)** | **clock·rate (95% HPD)** | **Genome-wide substitutions/year** |
| --- | --- | --- | --- | --- | --- | --- |
| Bayesian Skygrid | HKY | 100 | -12765·0534 (-12847·956, -12692·2473) | 398·2962 (187·2997, 663·2699) | 0·000807 (0·00034, 0·00135) | 0·5330235 |
| Bayesian Skyline | HKY | 200 | -15122·0646 (-15170·2677, -15074·2055) | 482·3089 (226·9919, 805·9951) | 0·000918 (0·000312, 0·00166) | 0·606339 |
| Bayesian Skyline | HKY | 100 | -15125·9168 (-15180·8875, -15079·4277) | 502·7905 (224·6944, 862·1201) | 0·000869 (0·000264, 0·00158) | 0·5739745 |
| Bayesian Skyline | GTR | 100 | -15127·5913 (-15175·8001, -15083·5638) * | 367·5269 (182·6824, 637·2581) | 0·000895 (0·000337, 0·00158) | 0·5911475 |
